# Early initiation of corticosteroids in patients hospitalized with COVID-19 not requiring intensive respiratory support: cohort study

**DOI:** 10.1101/2021.07.06.21259982

**Authors:** Kristina Crothers, Rian DeFaccio, Janet Tate, Patrick R. Alba, Matthew Goetz, Barbara Jones, Joseph T. King, Vincent Marconi, Michael E. Ohl, Christopher T. Rentsch, Maria C. Rodriguez-Barradas, Shahida Shahrir, Amy C. Justice, Kathleen M. Akgün, Veterans Aging Cohort Study Clinical COVID-19 Working Group

## Abstract

**Objectives:** To determine whether early oral or parenteral corticosteroids compared to no corticosteroids are associated with decreased mortality in patients hospitalized with coronavirus disease 2019 (COVID-19) who are not on intensive respiratory support (IRS) within 48 hours of admission.

**Design:** Observational cohort study

**Setting:** Nationwide cohort of patients receiving care in the Department of Veterans Affairs, a large integrated US national healthcare system

**Participants:** 9,058 patients admitted to a Veterans Affairs Medical Center between June 7, 2020-December 5, 2020 within 14-days after SARS-CoV-2 positive test; exclusion criteria include less than a 48 hour stay, receipt of prior systemic corticosteroids, and no indication of acute medical care for COVID-19.

**Main outcome measure:** 90-day all-cause mortality

**Results:** Of 9,058 total patients (95% men, median age 71 years, 27% black), 6,825 (75%) were not on IRS within 48 hours. Among the 3,025 patients on no oxygen, 598 (20%) received corticosteroids and 283 (9%) died; of 3,800 patients on low-flow nasal cannula oxygen (NC), 2,808 (74%) received corticosteroids and 514 (13%) died. In stratified, inverse probability weighted Cox proportional hazards models comparing those who did and did not receive corticosteroids, patients on no oxygen experienced an 89% increased risk for 90-day mortality (hazard ratio [HR] 1.89, 95% confidence interval [CI] 1.33 to 2.68); there was weak evidence of increased mortality among patients on NC (HR 1.21, 95% CI 0.94 to 1.57). Results were robust in subgroup analyses including restricting corticosteroids to dexamethasone, and in sensitivity analyses employing different modeling approaches.

**Conclusions:** In patients hospitalized with COVID-19, we found no evidence of a mortality benefit associated with early initiation of corticosteroids among those on no oxygen or NC in the first 48 hours, though there was evidence of potential harm. These real-world findings support that clinicians should consider withholding corticosteroids in these populations and further clinical trials may be warranted.

## INTRODUCTION

Corticosteroids have emerged as an effective therapy for critically ill patients with COVID-19. The United Kingdom RECOVERY trial demonstrated an overall 2.8% absolute decrease in mortality for patients treated with dexamethasone compared to usual care.^1^ When stratified by degree of respiratory support at randomization, the magnitude of benefit associated with dexamethasone was greater amongst those on invasive mechanical ventilation (IMV) versus supplemental oxygen (inclusive of non-invasive mechanical ventilation [NIV]); dexamethasone was not associated with significant mortality benefit amongst those not on oxygen.

Dissemination of these and other results led to nearly universal use of corticosteroids for COVID-19 patients receiving respiratory support, particularly more intensive respiratory support (IRS) such as high-flow nasal cannula (HFNC), NIV and IMV.^2-5^ Corticosteroids are also commonly used in patients not on IRS, despite being potentially less beneficial. Some studies investigating the association between corticosteroids and outcomes among a wider group of patients with COVID-19 – including a larger proportion not on IRS than in the RECOVERY trial – have found mixed results.^6-10^ Variability may be due to host factors including immune response to infection, heterogeneity in severity of illness and level of respiratory support, duration of infection, and use of different formulations of corticosteroids^8^ as well as residual confounding in observational studies. However, the findings may also suggest that certain subgroups of patients with COVID-19 benefit less from corticosteroids.

We sought to determine the association between corticosteroids and mortality using real-world clinical data from the Department of Veterans Affairs (VA), the largest integrated healthcare system in the United States. In a large, racially and geographically diverse, national cohort of hospitalized COVID-19 patients, we first assessed patterns of corticosteroid receipt. As nearly all patients on IRS received corticosteroids, we limited analysis of associations with 90-day mortality to those not on IRS. Because sicker patients are more likely to receive corticosteroids and more likely to die, we used propensity score weighting to account for confounding by indication.

## METHODS

### Study Design and Population

We conducted an observational cohort study using VA electronic health record (EHR) data. We identified an initial cohort of 12,455 patients with COVID-19 admitted to a VA hospital within 14 days after a positive polymerase chain reaction (PCR) or antigen test for SARS-CoV-2 between June 7, 2020 and December 5, 2020.^11,12^ Prior to June there was little use of corticosteroids in the first 48 hours. Index date was defined as date of presentation, including emergency room and any time spent under observation status prior to hospital admission if patients were not admitted directly. We determined hospital length of stay by concatenating consecutive episodes of care separated by less than 24 hours, with first episode of care on the index date identified as day one. Due to changes in COVID-19 incidence and treatment protocols over time, we divided the observation period into four phases: June 7-July 11; July 12-August 15; August 16-October 17; and October 18-December 5, 2020.

### Exclusions

From the initial 12,455 patients, we excluded 3,397. The most common reason was length of stay less than 48-hours (n=1,619; 40 deaths), as these patients had insufficient time to receive corticosteroids (Figure 1), followed by systemic corticosteroid exposure prior to index date. This was defined as any duration of corticosteroids within 14 days, or receipt of corticosteroids for ≥14 days within preceding 45 days (n=879). Within each phase, we restricted to facilities with at least 10 cases of COVID-19 and at least one corticosteroid prescription (N = 107 facilities) to have sufficient variation and number of events at each site. We excluded patients who were transferred from another acute care hospital (VA or non-VA) and who were likely incidentally-detected through screening prior to or at admission; this included patients who were not admitted to an acute medical care service or had no International Classification of Diseases, 10^th^ Revision (ICD-10) codes for COVID-19. We also excluded 101 patients prescribed hydroxychloroquine yielding a cohort of 9,058 patients.

**Figure 1.**
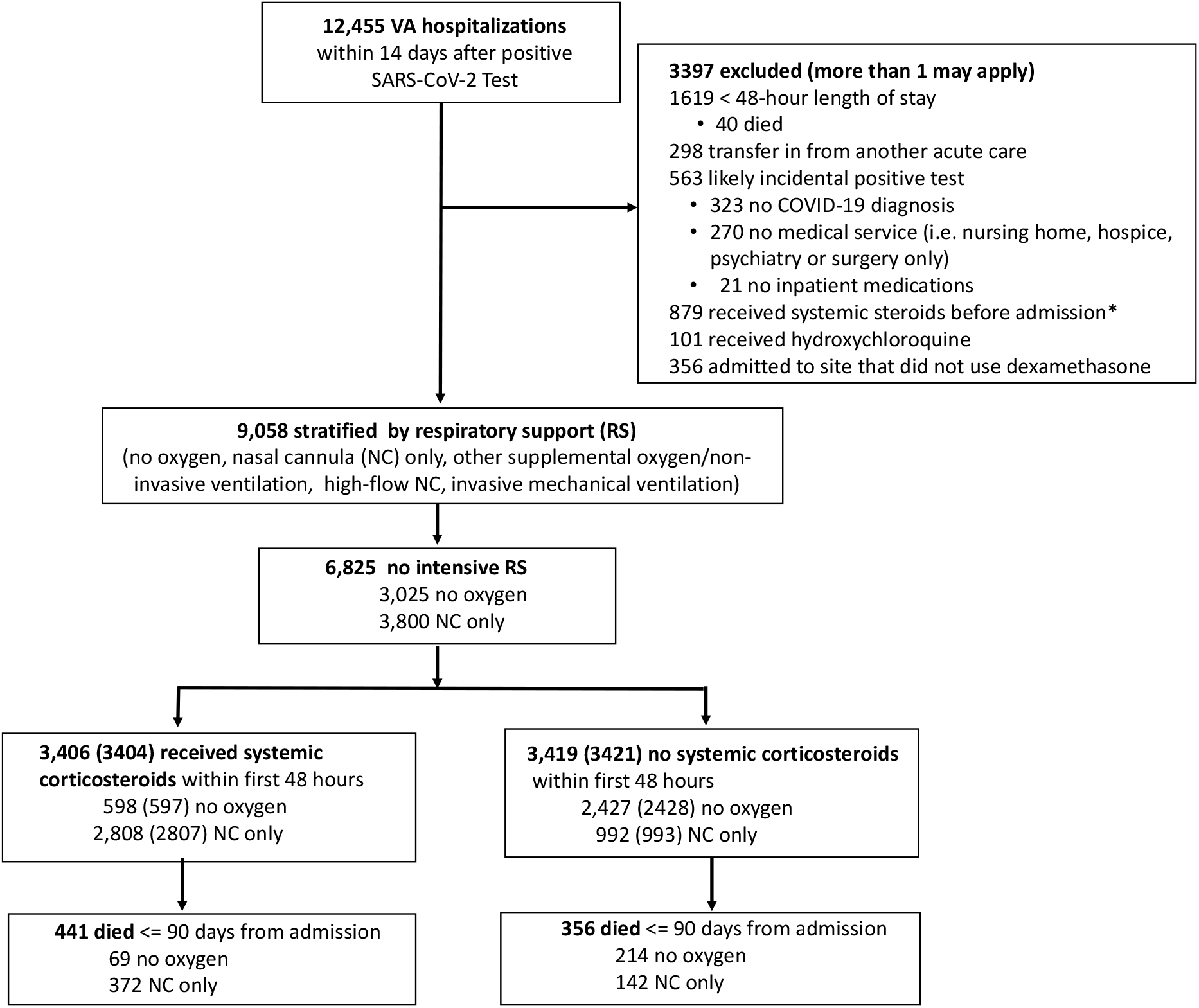
Derivation of Study Population.

### Respiratory Support

Next, we stratified patients by highest level of respiratory support during the initial 48 hours of hospitalization: 1) no oxygen support; 2) supplemental low-flow oxygen via nasal cannula (NC) that was not identified as a high-flow delivery device; 3) other supplemental oxygen/non-invasive ventilation (NIV), inclusive of oxygen by face mask, non-rebreather mask, NIV, or other forms of oxygen delivery not identifiable as low-flow NC or high-flow; 4) high-flow oxygen (abbreviated as high flow nasal cannula [HFNC]); and 5) invasive mechanical ventilation (IMV). When no evidence of oxygen supplementation was found, patients were classified as without oxygen (category 1). IMV was identified by structured data sources (ICD-10 procedure and Current Procedural Terminology [CPT] codes). Categories 2-4 were assessed from unstructured text notes using natural language processing (NLP) to identify key terms indicative of respiratory support. Schemas were developed iteratively with clinician review, including appropriate negation terms, based on snippets and note context. Patients on positive airway pressure (PAP) for sleep apnea without supplemental oxygen were classified as no oxygen. The NLP system was validated on 100 complete patient admissions, comprising 1,093 days reviewed. Fifty admissions were double annotated and adjudicated demonstrating Cohen’s kappa of 0.89. At the admission-level, receipt of any supplemental oxygen in categories 2-4 (NC, other/NIV and HFNC) was identified by the NLP system with a sensitivity, specificity, and positive predictive value of 100%, 77% and 94% respectively. When limited to the first 48 hours of admission, the system distinguished patients not on oxygen or on NC only from all other categories with 92% accuracy.

### Corticosteroid exposure

Corticosteroid administration (inclusive of dexamethasone, prednisone, prednisolone, methylprednisolone and/or hydrocortisone) was determined from bar code medication administration (BCMA) data (including intravenous). Patients who received at least one dose of an oral or parenteral corticosteroid within 48 hours after index date were considered exposed. We also determined the number of days during hospitalization that corticosteroids were administered. In each phase, we defined corticosteroid administration by site as low (<25^th^ percentile), medium (between 25-75% percentile), or high (>75^th^ percentile) based on the proportion in the sample receiving corticosteroids.

### Outcomes

The primary outcome was 90-day all-cause mortality, ascertained using inpatient records and VA death registry data to capture deaths outside of hospitalization. All hospitalizations occurred at least 90 days prior to the most recent mortality data. We also determined the proportion who initiated IMV or were administered vasopressors after the first 48 hours. Given low numbers of events, we did not conduct multivariable analyses for these outcomes.

### Covariates

We used EHR data to obtain demographics (age, race, ethnicity, sex), comorbidities, additional medications, and lab results, as well as to calculate the Charlson Comorbidity Index (CCI)^13^ and the Veterans Health Administration COVID-19 (VACO) Index.^14^ We focused on routinely collected laboratory tests that have been associated with increased mortality in COVID-19^15^ including albumin, liver function, lactate, white blood cell count, and creatinine (Table 2 and Supplemental Table 1). We selected the worst laboratory and vital signs values within the initial 48 hours. To account for potential effects of co-prescribed medications on COVID-19 outcomes, we also included use of remdesivir and prophylactic anticoagulants within the initial 48 hours.^12^ Intensive care unit (ICU) admission was determined using VA bedsection codes.^12,16^

### Statistical analysis

We initially compared COVID-19 patients by the five respiratory support categories using summary statistics (Table 1). Because nearly all patients on IMV or HFNC received corticosteroids; there was insufficient variability to allow generation of propensity score weights as discussed below. Category 3, “Other/NIV” respiratory support, was heterogenous with respect to support used and illness severity. For these reasons, as well as the greater clinical equipoise, we restricted our analysis to patients not on IRS (i.e., those on only NC or no oxygen support); from these, we further excluded 28 patients administered vasopressors in the first 48 hours from subsequent mortality analyses as they could have had an alternative indication for corticosteroids.

**TABLE 1:** Characteristics of patients stratified by highest oxygen support during first 48 hours of hospitalization for COVID-19.

|  | Oxygen Support first 48 hours |  |  |  |  |  |
| --- | --- | --- | --- | --- | --- | --- |
|  | Overall | None | Nasal Cannula | Other Supplemental<br>Oxygen/NIV | High Flow<br>Nasal Cannula | Invasive Mechanical<br>Ventilation |
| **Overall Cohort, n** | 9,058 | 3,025 | 3,800 | 813 | 1,194 | 226 |
| **Demographics** |  |  |  |  |  |  |
| **Age, n (%)** |  |  |  |  |  |  |
| <50 | 806 (9%) | 332 (11%) | 331 (9%) | 52 (6%) | 73 (6%) | 18 (8%) |
| 50-59 | 1,081 (12%) | 389 (13%) | 470 (12%) | 79 (10%) | 121 (10%) | 22 (10%) |
| 60-69 | 2,134 (24%) | 692 (23%) | 874 (23%) | 202 (25%) | 300 (25%) | 66 (29%) |
| 70-79 | 3,259 (36%) | 970 (32%) | 1,397 (37%) | 320 (39%) | 479 (40%) | 93 (41%) |
| 80+ | 1,778 (20%) | 642 (21%) | 728 (19%) | 160 (20%) | 221 (19%) | 27 (12%) |
| **Sex: Male, n (%)** | 8,606 (95%) | 2,858 (94%) | 3,594 (95%) | 775 (95%) | 1,163 (97%) | 216 (96%) |
| **Race, n (%)** |  |  |  |  |  |  |
| White, non-Hispanic | 4,989 (55%) | 1,563 (52%) | 2,121 (56%) | 480 (59%) | 704 (59%) | 121 (54%) |
| Black, non-Hispanic | 2,426 (27%) | 928 (31%) | 985 (26%) | 211 (26%) | 258 (22%) | 44 (19%) |
| Hispanic | 864 (10%) | 276 (9%) | 371 (10%) | 57 (7%) | 125 (10%) | 35 (15%) |
| Other | 491 (5%) | 152 (5%) | 208 (5%) | 46 (6%) | 70 (6%) | 15 (7%) |
| Unknown | 288 (3%) | 106 (4%) | 115 (3%) | 19 (2%) | 37 (3%) | 11 (5%) |
| **Phase (Admission Date, 2020), n (%)** |  |  |  |  |  |  |
| 1: June 7 - July 11 | 1,365 (15%) | 483 (16%) | 563 (15%) | 103 (13%) | 169 (14%) | 47 (21%) |
| 2: July 12 - Aug. 15 | 1,590 (18%) | 537 (18%) | 695 (18%) | 140 (17%) | 186 (16%) | 32 (14%) |
| 3: Aug. 16 - Oct. 17 | 1,908 (21%) | 668 (22%) | 792 (21%) | 163 (20%) | 229 (19%) | 56 (25%) |
| 4: Oct. 18 - Dec. 5 | 4,195 (46%) | 1,337 (44%) | 1,750 (46%) | 407 (50%) | 610 (51%) | 91 (40%) |
| **Selected Conditions** |  |  |  |  |  |  |
| **Dementia, n (%)** | 1,193 (13%) | 527 (17%) | 429 (11%) | 123 (15%) | 99 (8%) | 15 (7%) |
| **CHF, n (%)** | 1,886 (21%) | 571 (19%) | 810 (21%) | 233 (29%) | 233 (20%) | 39 (17%) |
| **COPD/Asthma, n (%)** | 2,715 (30%) | 729 (24%) | 1,251 (33%) | 296 (36%) | 377 (32%) | 62 (27%) |
| **Charlson Comorbidity Index, n (%)** |  |  |  |  |  |  |
| 0 | 1,706 (19%) | 634 (21%) | 698 (18%) | 98 (12%) | 233 (20%) | 43 (19%) |
| 1-2 | 2,868 (32%) | 940 (31%) | 1,216 (32%) | 219 (27%) | 409 (34%) | 84 (37%) |
| 3-4 | 2,145 (24%) | 660 (22%) | 940 (25%) | 224 (28%) | 274 (23%) | 47 (21%) |
| 5+ | 2,339 (26%) | 791 (26%) | 946 (25%) | 272 (33%) | 278 (23%) | 52 (23%) |
| **Medication Use** |  |  |  |  |  |  |
| **Steroid, Any Systemic, n (%)** |  |  |  |  |  |  |
| First 48 Hours | 5,165 (57%) | 598 (20%) | 2,808 (74%) | 468 (58%) | 1,083 (91%) | 208 (92%) |
| Later than 48 Hours | 870 (10%) | 363 (12%) | 337 (9%) | 115 (14%) | 43 (4%) | 12 (5%) |
| None | 3,023 (33%) | 2,064 (68%) | 655 (17%) | 230 (28%) | 68 (6%) | 6 (3%) |
| **Dexamethasone, n (%)** |  |  |  |  |  |  |
| First 48 Hours | 4,911 (54%) | 511 (17%) | 2,712 (71%) | 430 (53%) | 1,057 (89%) | 201 (89%) |

| Oxygen Support first 48 hours |  |  |  |  |  |  |
| --- | --- | --- | --- | --- | --- | --- |
|  | Overall | None | Nasal Cannula | Other Supplemental<br>Oxygen/NIV | High Flow<br>Nasal Cannula | Invasive Mechanical<br>Ventilation |
| Later than 48 Hours | 875 (10%) | 354 (12%) | 344 (9%) | 117 (14%) | 45 (4%) | 15 (7%) |
| None | 3,272 (36%) | 2,160 (71%) | 744 (20%) | 266 (33%) | 92 (8%) | 10 (4%) |
| **Remdesivir, n (%)** |  |  |  |  |  |  |
| First 48 Hours | 4,078 (45%) | 434 (14%) | 2,222 (58%) | 340 (42%) | 928 (78%) | 154 (68%) |
| Later than 48 Hours | 911 (10%) | 314 (10%) | 398 (10%) | 106 (13%) | 72 (6%) | 21 (9%) |
| None | 4,069 (45%) | 2,277 (75%) | 1,180 (31%) | 367 (45%) | 194 (16%) | 51 (23%) |
| **Prophylactic Anticoagulant, n (%)** |  |  |  |  |  |  |
| First 48 Hours | 5,020 (55%) | 1,606 (53%) | 2,180 (57%) | 438 (54%) | 666 (56%) | 130 (58%) |
| Later than 48 Hours | 549 (6%) | 175 (6%) | 210 (6%) | 53 (7%) | 94 (8%) | 17 (8%) |
| None | 3,489 (39%) | 1,244 (41%) | 1,410 (37%) | 322 (40%) | 434 (36%) | 79 (35%) |
| **Intensive Care, n (%)** |  |  |  |  |  |  |
| First 48 Hours | 2,006 (22%) | 278 (9%) | 583 (15%) | 203 (25%) | 735 (62%) | 207 (92%) |
| Later than 48 Hours | 676 (7%) | 124 (4%) | 352 (9%) | 76 (9%) | 113 (9%) | 11 (5%) |
| None | 6,376 (70%) | 2,623 (87%) | 2,865 (75%) | 534 (66%) | 346 (29%) | 8 (4%) |
| **Vasopressors, n (%)** |  |  |  |  |  |  |
| First 48 Hours | 188 ( 2.1%) | 9 (0.3%) | 19 ( 0.5%) | 11 ( 1.4%) | 27 ( 2.3%) | 122 (54.0%) |
| Later than 48 Hours | 539 ( 6.0%) | 47 (1.6%) | 169 ( 4.4%) | 44 ( 5.4%) | 217 (18.2%) | 62 (27.4%) |
| None | 8331 (92.0%) | 2,969 (98.1%) | 3612 (95.1%) | 758 (93.2%) | 950 (79.6%) | 42 (18.6%) |
| **Intubation, n (%)** |  |  |  |  |  |  |
| First 48 Hours | 226 (2%) | 0 (0%) | 0 (0%) | 0 (0%) | 0 (0%) | 226 (100%) |
| Later than 48 Hours | 492 (5%) | 49 (2%) | 161 (4%) | 46 (6%) | 236 (20%) | 0 (0%) |
| None | 8,340 (92%) | 2,976 (98%) | 3,639 (96%) | 767 (94%) | 958 (80%) | 0 (0%) |
| **Hospital Length of Stay, days,**<br>**median (interquartile range)** | 6 (4 – 12) | 5 (3 – 11) | 6 (4 – 10) | 6 (4 – 12) | 10 (6 – 17) | 16 (10 – 30) |
| **Mortality (unadjusted, cumulative**<br>**incidence)** |  |  |  |  |  |  |
| **30 Days, n (%)** | 1,151 (13%) | 184 (6%) | 375 (10%) | 124 (15%) | 360 (30%) | 108 (48%) |
| **60 Days, n (%)** | 1,413 (16%) | 247 (8%) | 477 (13%) | 164 (20%) | 404 (34%) | 121 (54%) |
| **90 Days, n (%)** | 1,512 (17%) | 283 (9%) | 514 (14%) | 175 (22%) | 416 (35%) | 124 (55%) |

In those not on IRS, we compared mortality by exposure to corticosteroids stratified by NC and overall. To account for confounding by indication, we generated propensity scores for the probability of receiving corticosteroids in the first 48 hours using logistic regression. Models included covariates thought to be associated with corticosteroid exposure and mortality: comorbidities, laboratory results, vital signs, site utilization patterns, co-medications and the 4 phases (Table 2 and Supplemental Table 1). We constructed inverse probability of treatment weights (IPTW) from propensity scores for each patient to create pseudo-populations with balanced distributions of covariates.^17^ There was generally very little missing data (<5%) so an explicit level for missingness was used for selected covariates. In our primary analysis, we used the average treatment effect (ATE) weights, reflecting the overall population from which the sample was taken. Stabilized weights were used and the ten patients with the most extreme high and low weights were trimmed from the analysis.^18^ We calculated standardized mean differences (SMD) between treatment groups and considered 0.2 or less as balanced. To compare differences in survival we generated IPTW Kaplan-Meier (KM) plots^19^ and Cox proportional hazards models estimating the ATE with days since index date as the time scale to provide formal hazard ratio (HR) risk estimates with confidence limits using a robust variance estimator. We included the VACO Index in outcome models to further account for residual confounding.^14^

**Table 2.**
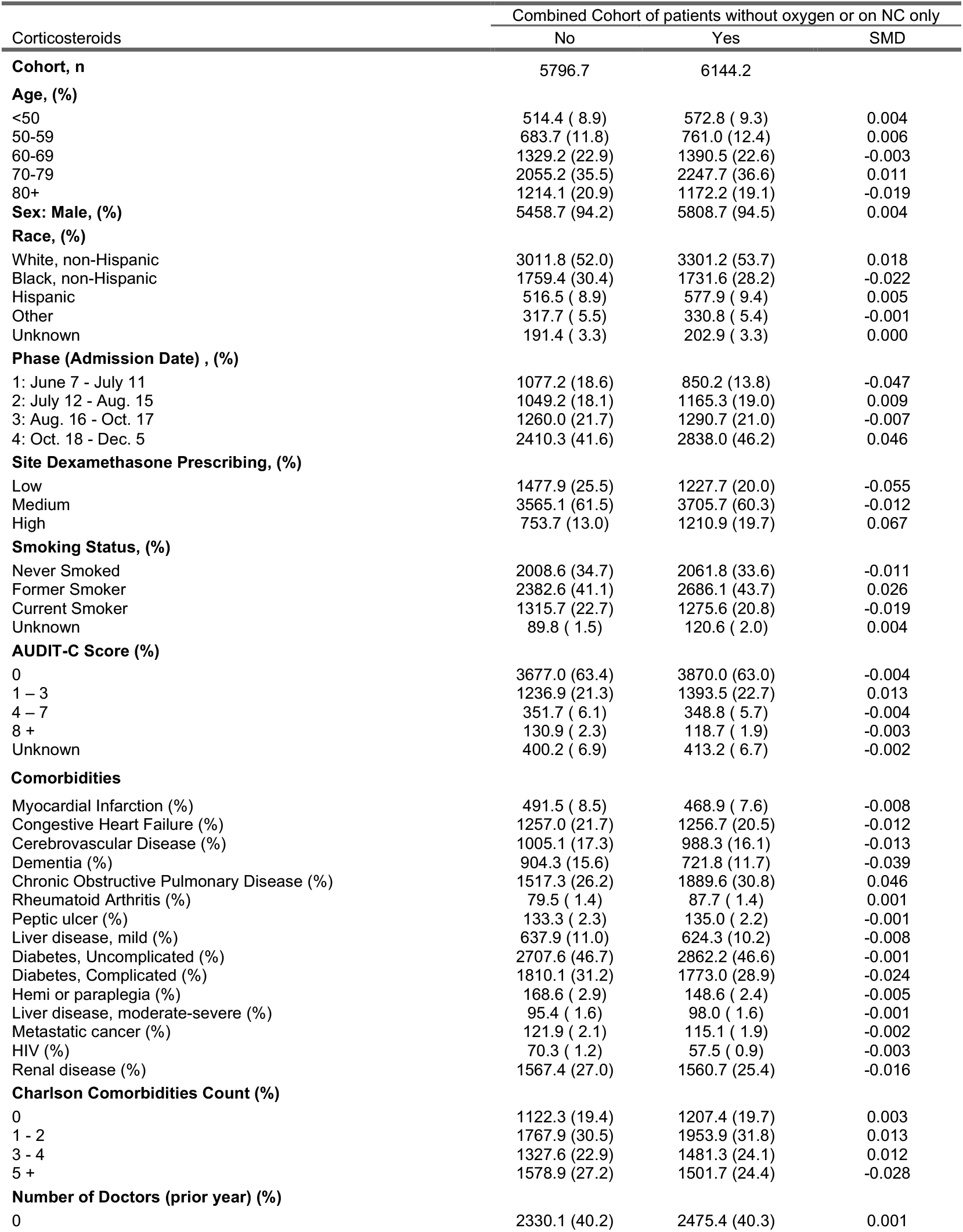
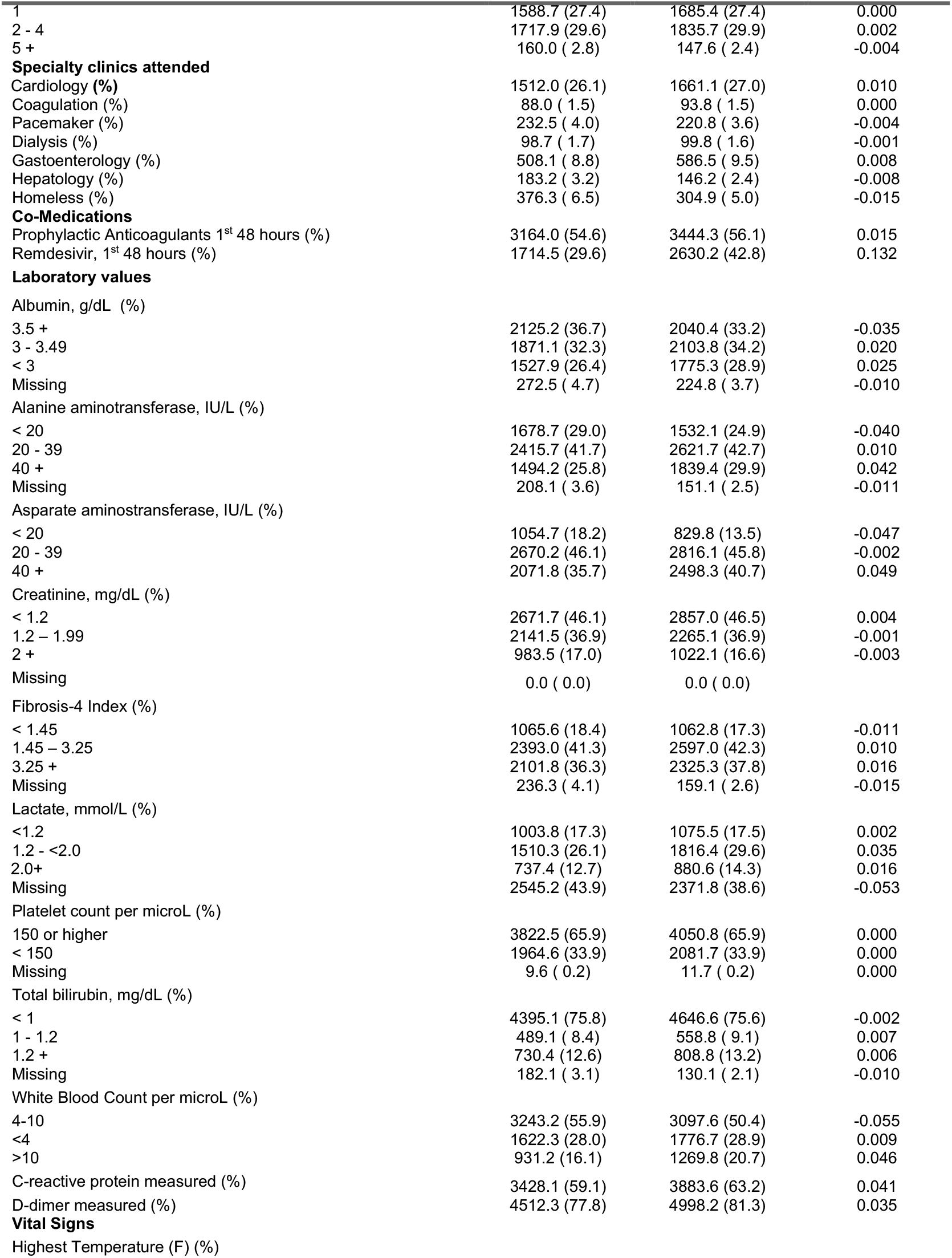
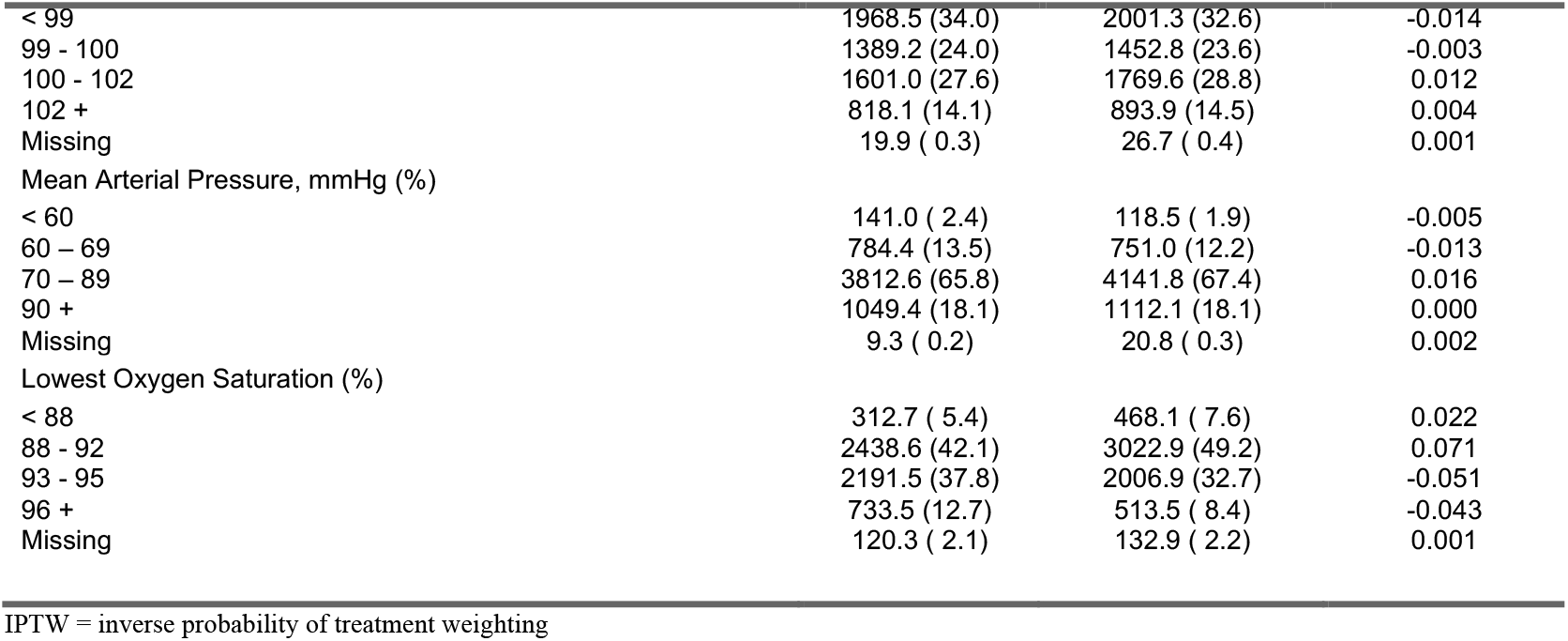
Characteristics of patients without oxygen or on NC after inverse probability of treatment weighting (IPTW) for estimating the average treatment effect in the total population (ATE models)

| Corticosteroids | Combined Cohort of patients without oxygen or on NC only |  |  |
| --- | --- | --- | --- |
|  | No | Yes | SMD |
| **Cohort, n** | 5796.7 | 6144.2 |  |
| **Age, (%)** |  |  |  |
| <50 | 514.4 ( 8.9) | 572.8 ( 9.3) | 0.004 |
| 50-59 | 683.7 (11.8) | 761.0 (12.4) | 0.006 |
| 60-69 | 1329.2 (22.9) | 1390.5 (22.6) | -0.003 |
| 70-79 | 2055.2 (35.5) | 2247.7 (36.6) | 0.011 |
| 80+ | 1214.1 (20.9) | 1172.2 (19.1) | -0.019 |
| **Sex: Male, (%)** | 5458.7 (94.2) | 5808.7 (94.5) | 0.004 |
| **Race, (%)** |  |  |  |
| White, non-Hispanic | 3011.8 (52.0) | 3301.2 (53.7) | 0.018 |
| Black, non-Hispanic | 1759.4 (30.4) | 1731.6 (28.2) | -0.022 |
| Hispanic | 516.5 ( 8.9) | 577.9 ( 9.4) | 0.005 |
| Other | 317.7 ( 5.5) | 330.8 ( 5.4) | -0.001 |
| Unknown | 191.4 ( 3.3) | 202.9 ( 3.3) | 0.000 |
| **Phase (Admission Date) , (%)** |  |  |  |
| 1: June 7 - July 11 | 1077.2 (18.6) | 850.2 (13.8) | -0.047 |
| 2: July 12 - Aug. 15 | 1049.2 (18.1) | 1165.3 (19.0) | 0.009 |
| 3: Aug. 16 - Oct. 17 | 1260.0 (21.7) | 1290.7 (21.0) | -0.007 |
| 4: Oct. 18 - Dec. 5 | 2410.3 (41.6) | 2838.0 (46.2) | 0.046 |
| **Site Dexamethasone Prescribing, (%)** |  |  |  |
| Low | 1477.9 (25.5) | 1227.7 (20.0) | -0.055 |
| Medium | 3565.1 (61.5) | 3705.7 (60.3) | -0.012 |
| High | 753.7 (13.0) | 1210.9 (19.7) | 0.067 |
| **Smoking Status, (%)** |  |  |  |
| Never Smoked | 2008.6 (34.7) | 2061.8 (33.6) | -0.011 |
| Former Smoker | 2382.6 (41.1) | 2686.1 (43.7) | 0.026 |
| Current Smoker | 1315.7 (22.7) | 1275.6 (20.8) | -0.019 |
| Unknown | 89.8 ( 1.5) | 120.6 ( 2.0) | 0.004 |
| **AUDIT-C Score (%)** |  |  |  |
| 0 | 3677.0 (63.4) | 3870.0 (63.0) | -0.004 |
| 1 – 3 | 1236.9 (21.3) | 1393.5 (22.7) | 0.013 |
| 4 – 7 | 351.7 ( 6.1) | 348.8 ( 5.7) | -0.004 |
| 8 + | 130.9 ( 2.3) | 118.7 ( 1.9) | -0.003 |
| Unknown | 400.2 ( 6.9) | 413.2 ( 6.7) | -0.002 |
| **Comorbidities** |  |  |  |
| Myocardial Infarction (%) | 491.5 ( 8.5) | 468.9 ( 7.6) | -0.008 |
| Congestive Heart Failure (%) | 1257.0 (21.7) | 1256.7 (20.5) | -0.012 |
| Cerebrovascular Disease (%) | 1005.1 (17.3) | 988.3 (16.1) | -0.013 |
| Dementia (%) | 904.3 (15.6) | 721.8 (11.7) | -0.039 |
| Chronic Obstructive Pulmonary Disease (%) | 1517.3 (26.2) | 1889.6 (30.8) | 0.046 |
| Rheumatoid Arthritis (%) | 79.5 ( 1.4) | 87.7 ( 1.4) | 0.001 |
| Peptic ulcer (%) | 133.3 ( 2.3) | 135.0 ( 2.2) | -0.001 |
| Liver disease, mild (%) | 637.9 (11.0) | 624.3 (10.2) | -0.008 |
| Diabetes, Uncomplicated (%) | 2707.6 (46.7) | 2862.2 (46.6) | -0.001 |
| Diabetes, Complicated (%) | 1810.1 (31.2) | 1773.0 (28.9) | -0.024 |
| Hemi or paraplegia (%) | 168.6 ( 2.9) | 148.6 ( 2.4) | -0.005 |
| Liver disease, moderate-severe (%) | 95.4 ( 1.6) | 98.0 ( 1.6) | -0.001 |
| Metastatic cancer (%) | 121.9 ( 2.1) | 115.1 ( 1.9) | -0.002 |
| HIV (%) | 70.3 ( 1.2) | 57.5 ( 0.9) | -0.003 |
| Renal disease (%) | 1567.4 (27.0) | 1560.7 (25.4) | -0.016 |
| **Charlson Comorbidities Count (%)** |  |  |  |
| 0 | 1122.3 (19.4) | 1207.4 (19.7) | 0.003 |
| 1 - 2 | 1767.9 (30.5) | 1953.9 (31.8) | 0.013 |
| 3 - 4 | 1327.6 (22.9) | 1481.3 (24.1) | 0.012 |
| 5 + | 1578.9 (27.2) | 1501.7 (24.4) | -0.028 |
| **Number of Doctors (prior year) (%)** |  |  |  |
| 0 | 2330.1 (40.2) | 2475.4 (40.3) | 0.001 |

|  | Combined Cohort of patients without oxygen or on NC only |  |  |
| --- | --- | --- | --- |
| Corticosteroids | No | Yes | SMD |
| 1 | 1588.7 (27.4) | 1685.4 (27.4) | 0.000 |
| 2 - 4 | 1717.9 (29.6) | 1835.7 (29.9) | 0.002 |
| 5 + | 160.0 ( 2.8) | 147.6 ( 2.4) | -0.004 |
| **Specialty clinics attended** |  |  |  |
| Cardiology (%) | 1512.0 (26.1) | 1661.1 (27.0) | 0.010 |
| Coagulation (%) | 88.0 ( 1.5) | 93.8 ( 1.5) | 0.000 |
| Pacemaker (%) | 232.5 ( 4.0) | 220.8 ( 3.6) | -0.004 |
| Dialysis (%) | 98.7 ( 1.7) | 99.8 ( 1.6) | -0.001 |
| Gastroenterology (%) | 508.1 ( 8.8) | 586.5 ( 9.5) | 0.008 |
| Hepatology (%) | 183.2 ( 3.2) | 146.2 ( 2.4) | -0.008 |
| Homeless (%) | 376.3 ( 6.5) | 304.9 ( 5.0) | -0.015 |
| **Co-Medications** |  |  |  |
| Prophylactic Anticoagulants 1<sup>st</sup> 48 hours (%) | 3164.0 (54.6) | 3444.3 (56.1) | 0.015 |
| Remdesivir, 1<sup>st</sup> 48 hours (%) | 1714.5 (29.6) | 2630.2 (42.8) | 0.132 |
| **Laboratory values** |  |  |  |
| Albumin, g/dL (%) |  |  |  |
| 3.5 + | 2125.2 (36.7) | 2040.4 (33.2) | -0.035 |
| 3 - 3.49 | 1871.1 (32.3) | 2103.8 (34.2) | 0.020 |
| < 3 | 1527.9 (26.4) | 1775.3 (28.9) | 0.025 |
| Missing | 272.5 ( 4.7) | 224.8 ( 3.7) | -0.010 |
| Alanine aminotransferase, IU/L (%) |  |  |  |
| < 20 | 1678.7 (29.0) | 1532.1 (24.9) | -0.040 |
| 20 - 39 | 2415.7 (41.7) | 2621.7 (42.7) | 0.010 |
| 40 + | 1494.2 (25.8) | 1839.4 (29.9) | 0.042 |
| Missing | 208.1 ( 3.6) | 151.1 ( 2.5) | -0.011 |
| Aspartate aminotransferase, IU/L (%) |  |  |  |
| < 20 | 1054.7 (18.2) | 829.8 (13.5) | -0.047 |
| 20 - 39 | 2670.2 (46.1) | 2816.1 (45.8) | -0.002 |
| 40 + | 2071.8 (35.7) | 2498.3 (40.7) | 0.049 |
| Creatinine, mg/dL (%) |  |  |  |
| < 1.2 | 2671.7 (46.1) | 2857.0 (46.5) | 0.004 |
| 1.2 – 1.99 | 2141.5 (36.9) | 2265.1 (36.9) | -0.001 |
| 2 + | 983.5 (17.0) | 1022.1 (16.6) | -0.003 |
| Missing | 0.0 ( 0.0) | 0.0 ( 0.0) |  |
| Fibrosis-4 Index (%) |  |  |  |
| < 1.45 | 1065.6 (18.4) | 1062.8 (17.3) | -0.011 |
| 1.45 – 3.25 | 2393.0 (41.3) | 2597.0 (42.3) | 0.010 |
| 3.25 + | 2101.8 (36.3) | 2325.3 (37.8) | 0.016 |
| Missing | 236.3 ( 4.1) | 159.1 ( 2.6) | -0.015 |
| Lactate, mmol/L (%) |  |  |  |
| <1.2 | 1003.8 (17.3) | 1075.5 (17.5) | 0.002 |
| 1.2 - <2.0 | 1510.3 (26.1) | 1816.4 (29.6) | 0.035 |
| 2.0+ | 737.4 (12.7) | 880.6 (14.3) | 0.016 |
| Missing | 2545.2 (43.9) | 2371.8 (38.6) | -0.053 |
| Platelet count per microL (%) |  |  |  |
| 150 or higher | 3822.5 (65.9) | 4050.8 (65.9) | 0.000 |
| < 150 | 1964.6 (33.9) | 2081.7 (33.9) | 0.000 |
| Missing | 9.6 ( 0.2) | 11.7 ( 0.2) | 0.000 |
| Total bilirubin, mg/dL (%) |  |  |  |
| < 1 | 4395.1 (75.8) | 4646.6 (75.6) | -0.002 |
| 1 - 1.2 | 489.1 ( 8.4) | 558.8 ( 9.1) | 0.007 |
| 1.2 + | 730.4 (12.6) | 808.8 (13.2) | 0.006 |
| Missing | 182.1 ( 3.1) | 130.1 ( 2.1) | -0.010 |
| White Blood Count per microL (%) |  |  |  |
| 4-10 | 3243.2 (55.9) | 3097.6 (50.4) | -0.055 |
| <4 | 1622.3 (28.0) | 1776.7 (28.9) | 0.009 |
| >10 | 931.2 (16.1) | 1269.8 (20.7) | 0.046 |
| C-reactive protein measured (%) | 3428.1 (59.1) | 3883.6 (63.2) | 0.041 |
| D-dimer measured (%) | 4512.3 (77.8) | 4998.2 (81.3) | 0.035 |
| **Vital Signs** |  |  |  |
| Highest Temperature (F) (%) |  |  |  |
|  | No | Yes | SMD |
| < 99 | 1968.5 (34.0) | 2001.3 (32.6) | -0.014 |
| 99 - 100 | 1389.2 (24.0) | 1452.8 (23.6) | -0.003 |
| 100 - 102 | 1601.0 (27.6) | 1769.6 (28.8) | 0.012 |
| 102 + | 818.1 (14.1) | 893.9 (14.5) | 0.004 |
| Missing | 19.9 ( 0.3) | 26.7 ( 0.4) | 0.001 |
| Mean Arterial Pressure, mmHg (%) |  |  |  |
| < 60 | 141.0 ( 2.4) | 118.5 ( 1.9) | -0.005 |
| 60 – 69 | 784.4 (13.5) | 751.0 (12.2) | -0.013 |
| 70 – 89 | 3812.6 (65.8) | 4141.8 (67.4) | 0.016 |
| 90 + | 1049.4 (18.1) | 1112.1 (18.1) | 0.000 |
| Missing | 9.3 ( 0.2) | 20.8 ( 0.3) | 0.002 |
| Lowest Oxygen Saturation (%) |  |  |  |
| < 88 | 312.7 ( 5.4) | 468.1 ( 7.6) | 0.022 |
| 88 - 92 | 2438.6 (42.1) | 3022.9 (49.2) | 0.071 |
| 93 - 95 | 2191.5 (37.8) | 2006.9 (32.7) | -0.051 |
| 96 + | 733.5 (12.7) | 513.5 ( 8.4) | -0.043 |
| Missing | 120.3 ( 2.1) | 132.9 ( 2.2) | 0.001 |

IPTW = inverse probability of treatment weighting

### Subgroup and sensitivity analyses

In primary analyses, we included all corticosteroid formulations; in subgroup analyses, we restricted to only dexamethasone. We also excluded patients admitted to the ICU within the first 48 hours, and restricted to patients age 70 and older. In sensitivity analyses, we evaluated the average treatment effect among the treated (ATT) population that received corticosteroids in weighted Cox proportional hazards models, and constructed unweighted, but multivariable adjusted models for all primary and subgroup analyses (Supplemental Tables 2 and 3).

Statistical analyses were performed using SAS 9.4 (SAS Institute, Cary, North Carolina, USA) and R version 4.0.4. Statistical significance was defined as p < 0.05. Our study was reviewed by the Institutional Review Boards of VA Puget Sound Health Care System, VA Connecticut Healthcare System and Yale University, and was deemed exempt. Study findings are reported as per the Strengthening the Reporting of Observational Studies in Epidemiology (STROBE) guidelines (Supplemental Table 4).

### Patient and public involvement

Patients and the public were not consulted during the design or conduct of this study.

## RESULTS

### Patient characteristics, corticosteroid exposure and respiratory support

Patients hospitalized during the four phases (1,328; 1,564; 1,857; 4,121) were mostly male (95%). Median age was 71 years (interquartile range [IQR] 62-77); 55% were non-Hispanic white, 27% non-Hispanic black, and 9% Hispanic (Table 1). Most patients (81%) were admitted within one day after positive SARS-CoV-2 test. More than half (56%) received corticosteroids (95% dexamethasone) within the first 48 hours of admission, increasing from 38% in phase 1 to 64% in phase 4. Concurrent remdesivir and prophylactic anticoagulants were more common in those who received corticosteroids (remdesivir 39% vs. 13%; anticoagulants 33% vs. 21%) than in those who did not.

When stratified by highest level of respiratory support provided in the first 48 hours of admission, most (77%) patients were on either no oxygen (34%) or NC only (43%) (Table 1). Corticosteroids were administered to 20% without oxygen, 74% of patients on NC, 58% on other supplemental oxygen/NIV, 91% on HFNC, and 92% on IMV. Use of corticosteroids increased over time, particularly in patients on NC (Figure 2). Overall, unadjusted 90-day mortality was 17%, and varied by level of respiratory support ranging from 9% in those without oxygen, 13% in patients on NC, and 56% in patients receiving IMV (Figure 2).

**Figure 2.**
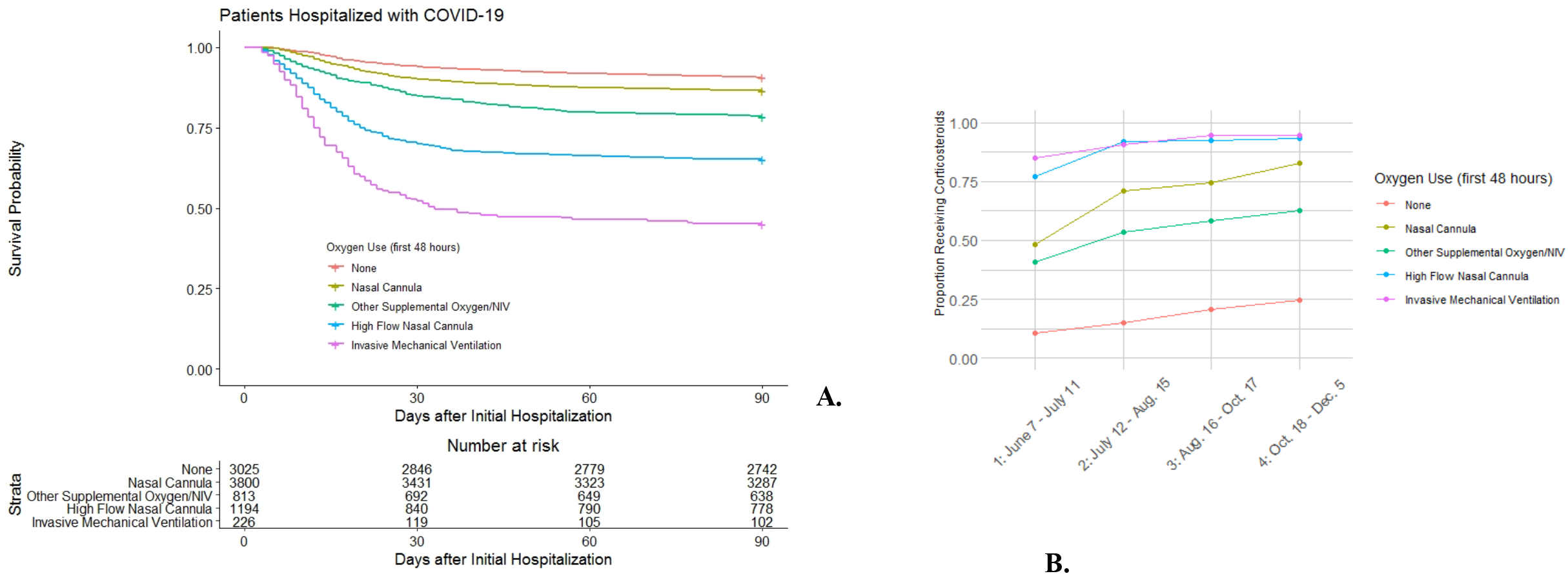
Unadjusted Kaplan Meier survival curves for 90-day mortality (A) and proportion exposed to corticosteroids by respiratory support level over time (B). RECOVERY Trial corticosteroid results issued in press release on 16 June 2020.

### Corticosteroids and mortality in patients not on IRS

Amongst patients not on IRS, the median duration of inpatient corticosteroids was 5 days (IQR 3-7) in patients without oxygen, and 6 days (IQR 4-9) in patients on NC. These were similar to hospital length of stay (Table 1). Only 80 (2.6%) and 79 (2.1%) patients, respectively, received one day of inpatient corticosteroids.

After propensity score weighting, our samples (pseudopopulations) were well-balanced, with covariates showing SMD generally <0.1; both overall and when split by no oxygen vs NC (Table 2 and Supplemental Table 1). Among patients without oxygen, weighted KM curves (Figure 3) show that those who received corticosteroids had higher mortality over 90-days than those who did not, with differences beginning to appear at 20-days after index date. In ATE estimates (Table 3), patients without oxygen who received corticosteroids had an 89% increased hazard of 90-day mortality (HR 1.89, 95% CI 1.33 to 2.68). Results were consistent in sensitivity analyses using ATT estimates as well as unweighted, multivariable adjusted Cox regression models (Supplemental Table 2).

**Table 3.** IPTW Cox Proportional Hazards Models for 90-day mortality associated with early corticosteroid exposure in patients hospitalized for COVID-19 among those not on IRS.

|  | **No oxygen supplementation** | **Nasal cannula** | **Combined group: no oxygen plus NC** |
| --- | --- | --- | --- |
|  | **HR (95% CI)** | **HR (95% CI)** | **HR (95% CI)** |
| **Primary analysis** | 1.89 (1.33 to 2.68) | 1.21 (0.94 to 1.57) | 1.61 (1.33 to 1.94) |
| **Subgroup analyses** |  |  |  |
| Restricted to dexamethasone | 2.03 (1.35 to 3.07) | 1.23 (0.89 to 1.71) | 1.57 (1.22 to 2.03) |
| Excluding patients admitted to ICU in initial 48 hours | 1.86 (1.22 to 2.83) | 1.30 (0.95 to 1.78) | 1.62 (1.28 to 2.06) |
| Restricted to patients age 70 and older | 1.91 (1.28 to 2.85) | 1.40 (1.03 to 1.92) | 1.65 (1.30 to 2.08) |

Models present the ATE (average treatment effect in entire population).
CI = confidence interval
HR = hazard ratio
IRS = intensive respiratory support

**Figure 3.**
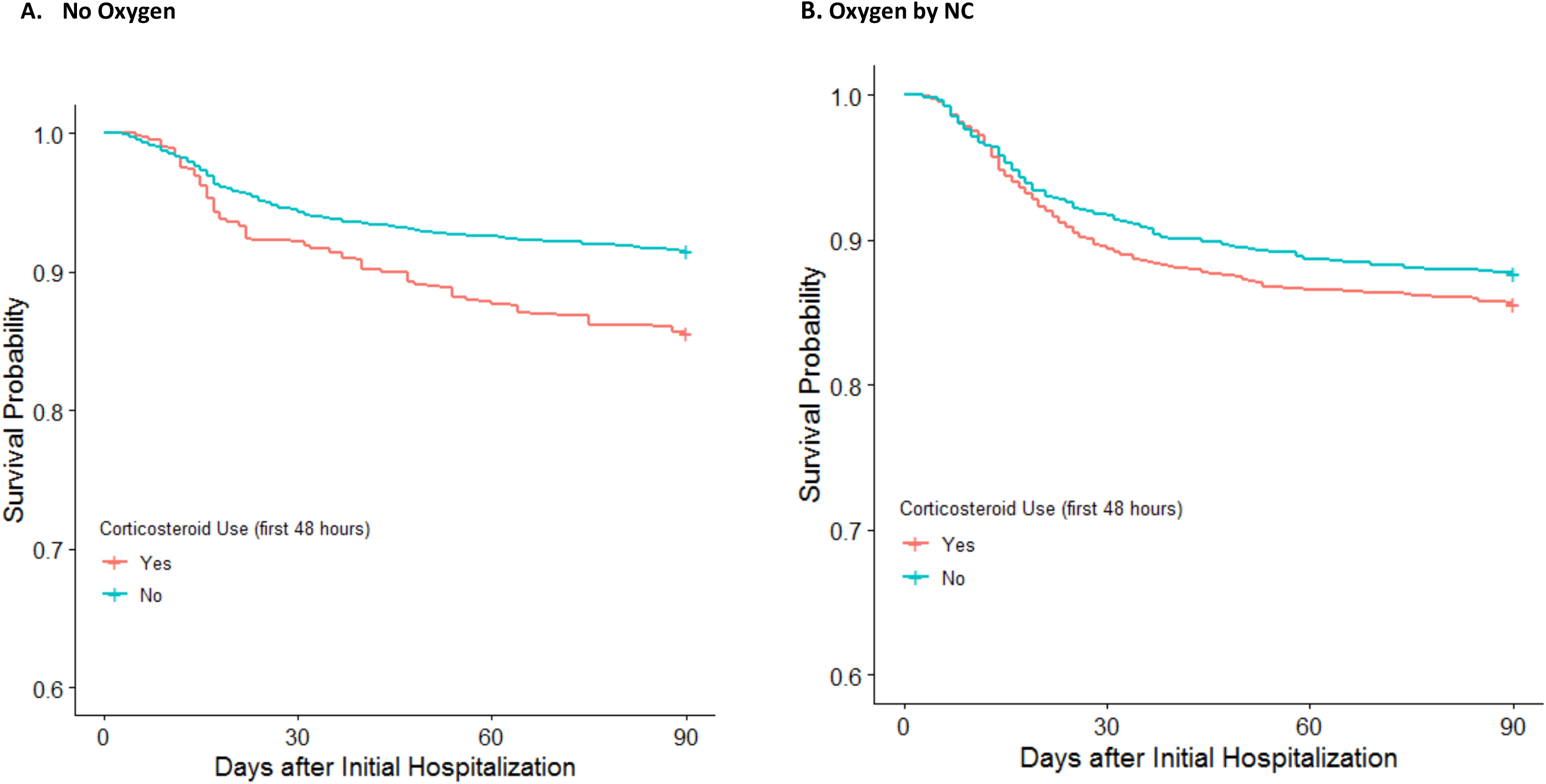
IPTW Kaplan Meier survival curves for 90-day mortality by corticosteroid use among those on no oxygen or NC. IPTW = Inverse probability of treatment weighting NC = nasal cannula

Similarly, patients on NC who received corticosteroids had higher 90-day mortality than those who did not, with weighted KM curves again diverging at 20-days (Figure 3). ATE estimates showed 21% increased mortality risk (HR 1.21, 05% CI 0.94 to 1.57) in patients who were on NC and received corticosteroids. HRs were similar in ATT estimates and multivariable Cox models (Supplemental Table 3). When combining patients on no oxygen or NC, corticosteroids were associated with an approximately 60% or more increased mortality risk.

### Subgroup analyses

Results were similar in subgroup analyses restricting corticosteroids to dexamethasone and excluding patients who were admitted to the ICU within the first 48 hours (Table 3). Limiting the sample to patients age 70 and older, mortality estimates associated with corticosteroids were slightly higher in the older population in those on NC. All findings were again consistent using ATT or multivariable Cox models (Supplemental Tables 2 and 3).

## DISCUSSION

In this analysis of a national cohort of hospitalized patients with COVID-19, receipt of corticosteroids was nearly universal among those on IRS and increasingly common among those not requiring it. Because corticosteroid receipt was nearly universal among those on IRS, we could not determine whether there was evidence of benefit. However, among patients not on oxygen in the first 48 hours of admission, corticosteroids initiated in this window were associated with increased 90-day mortality. Among those only on NC in the first 48 hours, corticosteroids were not associated with mortality benefit. Overall, when considering those not on IRS together, patients who received corticosteroids had on average a 60% or greater increase in mortality compared to those who did not receive corticosteroids.

Our findings were robust when assessed using several different approaches. Results were consistent using the ATE, reflecting the overall population from which the sample was taken, and using the ATT, reflecting the population of patients who received corticosteroids. They were also consistent controlling for potential confounders such as demographics, phase of the pandemic, site prescribing patterns, comorbidities, and laboratory values.^14^ Amongst patients age 70 and older, the mortality associated with corticosteroids appeared slightly greater in those on NC. Taken together, our findings suggest that the harms of corticosteroids – which become more apparent approximately 20 days after admission (Figure 3) – may outweigh benefits in hospitalized patients with COVID-19, particularly those who require no oxygen within 48 hours of admission. These findings may be under-estimated in studies where outcomes were assessed at 30-days and did not extend follow-up to 90-days.

From an implementation perspective, we found that uptake of corticosteroids for hospitalized COVID-19 patients was rapid. By mid-July 2020 most facilities had increased the proportion of patients administered corticosteroids to over 90% of those who were on HFNC or IMV within 48 hours of admission. Initiation of corticosteroids later during hospitalization was infrequent and decreased over time. Sites also increased use of corticosteroids for patients with less severe COVID-19, including those not on oxygen at the time of initiation (Figure 2). Patients not on IRS in the initial 48 hours represent the majority of COVID-19 admissions in the cohort; thus, our findings have important clinical implications on the potential unintended consequences of widespread corticosteroid adoption for COVID-19.

### Comparison with other evidence

While we cannot rule out residual confounding, results are consistent with the RECOVERY trial where the benefit of dexamethasone also diminished for patients not on IMV, and may have suggested possible harm in those not requiring oxygen.^1^ Recent meta-analyses and observational studies have also reported greater benefit of corticosteroids in severe compared to more mild acute respiratory distress syndrome (ARDS).^7,9^ However, these studies did not stratify patients further by level of respiratory support during the initial 48 hours of hospitalization, separating those who did and did not require IRS. Instead, clinical guidelines such as those issued by the National Institutes of Health recommend initiation of corticosteroids in hospitalized COVID-19 patients “on oxygen,” without further distinguishing between low-flow NC and forms of IRS.^20^ Our findings raise a note of caution when considering potential indication creep in the real world for use of corticosteroids in patients who do not have moderate-severe COVID-19 requiring IRS. Further randomized controlled trials of corticosteroids in patients with COVID-19 who are not on IRS may be warranted.

### Unanswered questions and future research

Even prior to COVID-19, the role of corticosteroids in lung infection has been uncertain. The impact of corticosteroids on patient outcomes has been inconsistent In other causes of ARDS including influenza and community-acquired pneumonia (CAP). In several viral pneumonias, corticosteroids were associated with greater risk of harm,^21-23^ although they appeared beneficial in the original severe acute respiratory syndrome.^7,24^ The net balance between benefit and harm of corticosteroids for pneumonia is not well understood and likely depends on multiple factors, including patient characteristics, heterogeneity in host response to infection, etiology of pneumonia, and time since onset of infection and ARDS.^25-28^ While corticosteroids may decrease host inflammatory response with potential modulating effects to lung injury, they may also have harmful side effects or unintended consequences on adaptive immune responses that may be important to resolution of infection. We saw potentially an increased risk of mortality associated with corticosteroids in patients on NC who were over age 70. Whether this is due to differences in immune responses in older individuals and/or greater risk of unintended consequences from corticosteroids, such as secondary infections, requires further study.

There remain additional unanswered questions regarding the timing of corticosteroids for patients hospitalized with COVID-19, particularly those not on IRS. When corticosteroids are initiated during the course of infection is likely important. For some patients, initiation within 48 hours of hospitalization may be too early in the course of disease and could impair viral clearance.^29^ While the majority of patients had positive SARS-CoV-2 testing within one day prior to hospitalization in our cohort, we do not know how long symptoms preceded a patient seeking medical attention and receiving SARS-CoV-2 testing. The optimal formulation, dose and duration of corticosteroids also remain unclear, and if these should vary depending on severity of COVID-19.^2-4,29^

### Strengths and weaknesses of this study

There are several limitations to our study. First, the study was observational. While we used detailed clinical data that included measures reflecting illness severity in a large population that demonstrated excellent balance in propensity for treatment, residual confounding for severity of illness could have contributed to greater mortality in those exposed to corticosteroids. In addition, the effects associated with initial corticosteroids are difficult to disentangle from other therapies that may have been concomitantly received. Some laboratory results used as covariates could have occurred after corticosteroid exposure, as both were ascertained within 48 hours. However, this approach allowed an equal time window to detect worst results in patients exposed and unexposed to corticosteroids and the impact of corticosteroids on acute laboratory results was likely limited. Although respiratory support algorithms were manually reviewed and validated, some misclassification may have occurred. Nonetheless, the substantial separation in Kaplan-Meier mortality curves with increasing mortality with greater respiratory support supports the validity of this variable. We also cannot rule out that clinicians were aware of another indication for corticosteroids beyond COVID-19 in patients who were not on oxygen or were on NC, such as airway inflammation from obstructive lung disease. However, we excluded those on steroids prior to admission and patients on vasopressors within the initial 48 hours, as these patients were more likely have an alternative indication for steroids. Further, we did not assess dose of corticosteroids or calculate total duration of exposure by including discharge medications after index hospitalization. Most patients had an average length of inpatient corticosteroid treatment that matched their hospital length of stay, and very few received only one dose of a corticosteroid (<3%). Finally, our cohort consisted predominantly of male Veterans, but had excellent racial and geographic variability.

### Conclusions and implications

In summary, we found that in patients with COVID-19 not on IRS in the first 48 hours of hospitalization, early corticosteroids were associated with increased 90-day mortality. Further, we failed to confirm a benefit of early corticosteroids to 90-day mortality in those on NC oxygen. These results were consistent in different analytic approaches that used real-world evidence with detailed clinical data in a large population to control for confounding, and remained robust in a variety of subgroup and sensitivity analyses. Our findings raise the possibility that harms of corticosteroids initiated within 48 hours of hospitalization may outweigh benefits in some patients with COVID-19 and less severe respiratory compromise, particularly in those who not on oxygen within 48 hours of hospital admission. Given the trend towards increasing uptake of corticosteroids in these populations, the real world evidence presented here highlights the non-beneficial and potentially harmful indication creep in the use of corticosteroids in hospitalized COVID-19 patients not on IRS. Clinicians should consider withholding corticosteroids in these populations and further clinical trials may be warranted.

## Data Availability

Owing to US Department of Veterans Affairs (VA) regulations and our ethics agreements, the analytic data sets used for this study are not permitted to leave the VA firewall without a data use agreement. This limitation is consistent with other studies based on VA data. However, VA data are made freely available to researchers with an approved VA study protocol. For more information, please visit https://www.virec.research.va.gov or contact the VA Information Resource Center at.

