## Supplemental Tables for "Early initiation of corticosteroids in patients hospitalized with COVID-19 not requiring intensive respiratory support: cohort study"

**Data supplement**

**Supplemental Table 1. Unweighted and propensity weighted pseudo populations**

**A. Unweighted population**

| Corticosteroids | No Oxygen | | | Nasal Canula | | | Combined Cohort | | |
| --- | --- | --- | --- | --- | --- | --- | --- | --- | --- |
|  | No | Yes | SMD | No | Yes | SMD | No | Yes | SMD |
| **Cohort, n** | 2421 | 595 |  | 988 | 2793 |  | 3409 | 3388 |  |
| **Age, (%)** |  |  |  |  |  |  |  |  |  |
| <50 | 273 (11.3) | 59 ( 9.9) | -0.014 | 70 ( 7.1) | 261 ( 9.3) | 0.023 | 343 (10.1) | 320 ( 9.4) | -0.006 |
| 50-59 | 296 (12.2) | 90 (15.1) | 0.029 | 125 (12.7) | 344 (12.3) | -0.003 | 421 (12.3) | 434 (12.8) | 0.005 |
| 60-69 | 551 (22.8) | 138 (23.2) | 0.004 | 222 (22.5) | 647 (23.2) | 0.007 | 773 (22.7) | 785 (23.2) | 0.005 |
| 70-79 | 772 (31.9) | 195 (32.8) | 0.009 | 341 (34.5) | 1048 (37.5) | 0.030 | 1113 (32.6) | 1243 (36.7) | 0.040 |
| 80+ | 529 (21.9) | 113 (19.0) | -0.029 | 230 (23.3) | 493 (17.7) | -0.056 | 759 (22.3) | 606 (17.9) | -0.044 |
| **Sex: Male, (%)** | 2287 (94.5) | 562 (94.5) | 0.000 | 931 (94.2) | 2644 (94.7) | 0.004 | 3218 (94.4) | 3206 (94.6) | 0.002 |
| **Race, (%)** |  |  |  |  |  |  |  |  |  |
| White, non-Hispanic | 1240 (51.2) | 321 (53.9) | 0.027 | 526 (53.2) | 1589 (56.9) | 0.037 | 1766 (51.8) | 1910 (56.4) | 0.046 |
| Black, non-Hispanic | 765 (31.6) | 159 (26.7) | -0.049 | 299 (30.3) | 678 (24.3) | -0.060 | 1064 (31.2) | 837 (24.7) | -0.065 |
| Hispanic | 213 ( 8.8) | 61 (10.3) | 0.015 | 78 ( 7.9) | 290 (10.4) | 0.025 | 291 ( 8.5) | 351 (10.4) | 0.018 |
| Other | 123 ( 5.1) | 28 ( 4.7) | -0.004 | 55 ( 5.6) | 152 ( 5.4) | -0.001 | 178 ( 5.2) | 180 ( 5.3) | 0.001 |
| Unknown | 80 ( 3.3) | 26 ( 4.4) | 0.011 | 30 ( 3.0) | 84 ( 3.0) | 0.000 | 110 ( 3.2) | 110 ( 3.2) | 0.000 |
| **Phase (Admission Date) , (%)** |  |  |  |  |  |  |  |  |  |
| 1: June 7 - July 11 | 430 (17.8) | 51 ( 8.6) | -0.092 | 289 (29.3) | 270 ( 9.7) | -0.196 | 719 (21.1) | 321 ( 9.5) | -0.116 |
| 2: July 12 - Aug. 15 | 456 (18.8) | 79 (13.3) | -0.056 | 201 (20.3) | 490 (17.5) | -0.028 | 657 (19.3) | 569 (16.8) | -0.025 |
| 3: Aug. 16 - Oct. 17 | 525 (21.7) | 139 (23.4) | 0.017 | 201 (20.3) | 587 (21.0) | 0.007 | 726 (21.3) | 726 (21.4) | 0.001 |
| 4: Oct. 18 - Dec. 5 | 1010 (41.7) | 326 (54.8) | 0.131 | 297 (30.1) | 1446 (51.8) | 0.217 | 1307 (38.3) | 1772 (52.3) | 0.140 |
| **Site Dexamethasone Prescribing, (%)** |  |  |  |  |  |  |  |  |  |
| Low | 690 (28.5) | 84 (14.1) | -0.144 | 295 (29.9) | 421 (15.1) | -0.148 | 985 (28.9) | 505 (14.9) | -0.140 |
| Medium | 1473 (60.8) | 358 (60.2) | -0.007 | 572 (57.9) | 1665 (59.6) | 0.017 | 2045 (60.0) | 2023 (59.7) | -0.003 |
| High | 258 (10.7) | 153 (25.7) | 0.151 | 121 (12.2) | 707 (25.3) | 0.131 | 379 (11.1) | 860 (25.4) | 0.143 |
| **Smoking Status, (%)** |  |  |  |  |  |  |  |  |  |
| Never Smoked | 842 (34.8) | 203 (34.1) | -0.007 | 337 (34.1) | 939 (33.6) | -0.005 | 1179 (34.6) | 1142 (33.7) | -0.009 |
| Former Smoker | 908 (37.5) | 269 (45.2) | 0.077 | 411 (41.6) | 1305 (46.7) | 0.051 | 1319 (38.7) | 1574 (46.5) | 0.078 |
| Current Smoker | 626 (25.9) | 112 (18.8) | -0.070 | 211 (21.4) | 506 (18.1) | -0.032 | 837 (24.6) | 618 (18.2) | -0.063 |
| Unknown | 45 ( 1.9) | 11 ( 1.8) | 0.000 | 29 ( 2.9) | 43 ( 1.5) | -0.014 | 74 ( 2.2) | 54 ( 1.6) | -0.006 |
| **AUDIT-C Score (%)** |  |  |  |  |  |  |  |  |  |
| 0 | 1489 (61.5) | 369 (62.0) | 0.005 | 645 (65.3) | 1767 (63.3) | -0.020 | 2134 (62.6) | 2136 (63.0) | 0.004 |
| 1 – 3 | 481 (19.9) | 132 (22.2) | 0.023 | 204 (20.6) | 674 (24.1) | 0.035 | 685 (20.1) | 806 (23.8) | 0.037 |
| 4 – 7 | 167 ( 6.9) | 34 ( 5.7) | -0.012 | 39 ( 3.9) | 147 ( 5.3) | 0.013 | 206 ( 6.0) | 181 ( 5.3) | -0.007 |
| 8 + | 81 ( 3.3) | 15 ( 2.5) | -0.008 | 20 ( 2.0) | 43 ( 1.5) | -0.005 | 101 ( 3.0) | 58 ( 1.7) | -0.013 |
| Unknown | 203 ( 8.4) | 45 ( 7.6) | -0.008 | 80 ( 8.1) | 162 ( 5.8) | -0.023 | 283 ( 8.3) | 207 ( 6.1) | -0.022 |
| **Comorbidities** |  |  |  |  |  |  |  |  |  |
| Myocardial Infarction (%) | 208 ( 8.6) | 36 ( 6.1) | -0.025 | 89 ( 9.0) | 216 ( 7.7) | -0.013 | 297 ( 8.7) | 252 ( 7.4) | -0.013 |
| Congestive Heart Failure (%) | 468 (19.3) | 102 (17.1) | -0.022 | 271 (27.4) | 534 (19.1) | -0.083 | 739 (21.7) | 636 (18.8) | -0.029 |
| Cerebrovascular Disease (%) | 461 (19.0) | 79 (13.3) | -0.058 | 188 (19.0) | 418 (15.0) | -0.041 | 649 (19.0) | 497 (14.7) | -0.044 |
| Dementia (%) | 467 (19.3) | 59 ( 9.9) | -0.094 | 172 (17.4) | 252 ( 9.0) | -0.084 | 639 (18.7) | 311 ( 9.2) | -0.096 |
| Chronic Obstructive Pulmonary Disease (%) | 546 (22.6) | 168 (28.2) | 0.057 | 306 (31.0) | 923 (33.0) | 0.021 | 852 (25.0) | 1091 (32.2) | 0.072 |
| Rheumatoid Arthritis (%) | 34 ( 1.4) | 9 ( 1.5) | 0.001 | 14 ( 1.4) | 49 ( 1.8) | 0.003 | 48 ( 1.4) | 58 ( 1.7) | 0.003 |
| Peptic ulcer (%) | 72 ( 3.0) | 9 ( 1.5) | -0.015 | 25 ( 2.5) | 55 ( 2.0) | -0.006 | 97 ( 2.8) | 64 ( 1.9) | -0.010 |
| Liver disease, mild (%) | 335 (13.8) | 66 (11.1) | -0.027 | 100 (10.1) | 265 ( 9.5) | -0.006 | 435 (12.8) | 331 ( 9.8) | -0.030 |
| Diabetes, Uncomplicated (%) | 1054 (43.5) | 278 (46.7) | 0.032 | 474 (48.0) | 1369 (49.0) | 0.010 | 1528 (44.8) | 1647 (48.6) | 0.038 |
| Diabetes, Complicated (%) | 704 (29.1) | 168 (28.2) | -0.008 | 318 (32.2) | 819 (29.3) | -0.029 | 1022 (30.0) | 987 (29.1) | -0.008 |
| Hemi or paraplegia (%) | 84 ( 3.5) | 10 ( 1.7) | -0.018 | 36 ( 3.6) | 52 ( 1.9) | -0.018 | 120 ( 3.5) | 62 ( 1.8) | -0.017 |
| Liver disease, moderate-severe (%) | 56 ( 2.3) | 9 ( 1.5) | -0.008 | 13 ( 1.3) | 32 ( 1.1) | -0.002 | 69 ( 2.0) | 41 ( 1.2) | -0.008 |
| Metastatic cancer (%) | 56 ( 2.3) | 6 ( 1.0) | -0.013 | 16 ( 1.6) | 46 ( 1.6) | 0.000 | 72 ( 2.1) | 52 ( 1.5) | -0.006 |
| HIV (%) | 32 ( 1.3) | 10 ( 1.7) | 0.004 | 13 ( 1.3) | 20 ( 0.7) | -0.006 | 45 ( 1.3) | 30 ( 0.9) | -0.004 |
| Renal disease (%) | 610 (25.2) | 140 (23.5) | -0.017 | 284 (28.7) | 656 (23.5) | -0.053 | 894 (26.2) | 796 (23.5) | -0.027 |
| **Charlson Comorbidities Count (%)** |  |  |  |  |  |  |  |  |  |
| 0 | 499 (20.6) | 134 (22.5) | 0.019 | 179 (18.1) | 518 (18.5) | 0.004 | 678 (19.9) | 652 (19.2) | -0.006 |
| 1 - 2 | 737 (30.4) | 200 (33.6) | 0.032 | 278 (28.1) | 934 (33.4) | 0.053 | 1015 (29.8) | 1134 (33.5) | 0.037 |
| 3 - 4 | 529 (21.9) | 131 (22.0) | 0.002 | 235 (23.8) | 702 (25.1) | 0.013 | 764 (22.4) | 833 (24.6) | 0.022 |
| 5 + | 656 (27.1) | 130 (21.8) | -0.052 | 296 (30.0) | 639 (22.9) | -0.071 | 952 (27.9) | 769 (22.7) | -0.052 |
| **Number of Doctors (prior year) (%)** |  |  |  |  |  |  |  |  |  |
| 0 | 1057 (43.7) | 250 (42.0) | -0.016 | 395 (40.0) | 1075 (38.5) | -0.015 | 1452 (42.6) | 1325 (39.1) | -0.035 |
| 1 | 653 (27.0) | 182 (30.6) | 0.036 | 246 (24.9) | 780 (27.9) | 0.030 | 899 (26.4) | 962 (28.4) | 0.020 |
| 2 - 4 | 645 (26.6) | 155 (26.1) | -0.006 | 316 (32.0) | 879 (31.5) | -0.005 | 961 (28.2) | 1034 (30.5) | 0.023 |
| 5 + | 66 ( 2.7) | 8 ( 1.3) | -0.014 | 31 ( 3.1) | 59 ( 2.1) | -0.010 | 97 ( 2.8) | 67 ( 2.0) | -0.009 |
| **Specialty clinics attended**  Cardiology **(%)** | 580 (24.0) | 147 (24.7) | 0.007 | 296 (30.0) | 759 (27.2) | -0.028 | 876 (25.7) | 906 (26.7) | 0.010 |
| Coagulation (%) | 38 ( 1.6) | 13 ( 2.2) | 0.006 | 21 ( 2.1) | 31 ( 1.1) | -0.010 | 59 ( 1.7) | 44 ( 1.3) | -0.004 |
| Pacemaker (%) | 93 ( 3.8) | 20 ( 3.4) | -0.005 | 48 ( 4.9) | 70 ( 2.5) | -0.024 | 141 ( 4.1) | 90 ( 2.7) | -0.015 |
| Dialysis (%) | 32 ( 1.3) | 5 ( 0.8) | -0.005 | 32 ( 3.2) | 33 ( 1.2) | -0.021 | 64 ( 1.9) | 38 ( 1.1) | -0.008 |
| Gastoenterology (%) | 203 ( 8.4) | 61 (10.3) | 0.019 | 86 ( 8.7) | 292 (10.5) | 0.018 | 289 ( 8.5) | 353 (10.4) | 0.019 |
| Hepatology (%) | 97 ( 4.0) | 10 ( 1.7) | -0.023 | 30 ( 3.0) | 64 ( 2.3) | -0.007 | 127 ( 3.7) | 74 ( 2.2) | -0.015 |
| Homeless (%) | 199 ( 8.2) | 30 ( 5.0) | -0.032 | 58 ( 5.9) | 93 ( 3.3) | -0.025 | 257 ( 7.5) | 123 ( 3.6) | -0.039 |
| Prophylactic Anticoagulants | 1267 (52.3) | 332 (55.8) | 0.035 | 523 (52.9) | 1643 (58.8) | 0.059 | 1790 (52.5) | 1975 (58.3) | 0.058 |
| **Co-medications** |  |  |  |  |  |  |  |  |  |
| Remdesivir, 1^st^ 48 hours (%) | 174 ( 7.2) | 257 (43.2) | 0.360 | 211 (21.4) | 2001 (71.6) | 0.503 | 385 (11.3) | 2258 (66.6) | 0.554 |
| **Laboratory Results** |  |  |  |  |  |  |  |  |  |
| Albumin, g/dL (%) |  |  |  |  |  |  |  |  |  |
| 3.5 + | 1050 (43.4) | 218 (36.6) | -0.067 | 305 (30.9) | 781 (28.0) | -0.029 | 1355 (39.7) | 999 (29.5) | -0.103 |
| 3 - 3.49 | 719 (29.7) | 203 (34.1) | 0.044 | 344 (34.8) | 1005 (36.0) | 0.012 | 1063 (31.2) | 1208 (35.7) | 0.045 |
| < 3 | 478 (19.7) | 151 (25.4) | 0.056 | 285 (28.8) | 928 (33.2) | 0.044 | 763 (22.4) | 1079 (31.8) | 0.095 |
| Missing | 174 ( 7.2) | 23 ( 3.9) | -0.033 | 54 ( 5.5) | 79 ( 2.8) | -0.026 | 228 ( 6.7) | 102 ( 3.0) | -0.037 |
| Alanine aminotransferase, IU/L (%) |  |  |  |  |  |  |  |  |  |
| < 20 | 784 (32.4) | 148 (24.9) | -0.075 | 290 (29.4) | 578 (20.7) | -0.087 | 1074 (31.5) | 726 (21.4) | -0.101 |
| 20 - 39 | 938 (38.7) | 242 (40.7) | 0.019 | 418 (42.3) | 1201 (43.0) | 0.007 | 1356 (39.8) | 1443 (42.6) | 0.028 |
| 40 + | 533 (22.0) | 199 (33.4) | 0.114 | 227 (23.0) | 977 (35.0) | 0.120 | 760 (22.3) | 1176 (34.7) | 0.124 |
| Missing | 166 ( 6.9) | 6 ( 1.0) | -0.058 | 53 ( 5.4) | 37 ( 1.3) | -0.040 | 219 ( 6.4) | 43 ( 1.3) | -0.052 |
| Asparate aminostransferase, IU/L (%) |  |  |  |  |  |  |  |  |  |
| < 20 | 654 (27.0) | 65 (10.9) | -0.161 | 199 (20.1) | 218 ( 7.8) | -0.123 | 853 (25.0) | 283 ( 8.4) | -0.167 |
| 20 - 39 | 1098 (45.4) | 279 (46.9) | 0.015 | 444 (44.9) | 1218 (43.6) | -0.013 | 1542 (45.2) | 1497 (44.2) | -0.010 |
| 40 + | 669 (27.6) | 251 (42.2) | 0.146 | 345 (34.9) | 1357 (48.6) | 0.137 | 1014 (29.7) | 1608 (47.5) | 0.177 |
| Creatinine, mg/dL (%) |  |  |  |  |  |  |  |  |  |
| < 1.2 | 1223 (50.5) | 277 (46.6) | -0.040 | 423 (42.8) | 1324 (47.4) | 0.046 | 1646 (48.3) | 1601 (47.3) | -0.010 |
| 1.2 – 1.99 | 797 (32.9) | 244 (41.0) | 0.081 | 374 (37.9) | 1048 (37.5) | -0.003 | 1171 (34.4) | 1292 (38.1) | 0.038 |
| 2 + | 360 (14.9) | 74 (12.4) | -0.024 | 186 (18.8) | 421 (15.1) | -0.038 | 546 (16.0) | 495 (14.6) | -0.014 |
| Missing | 41 ( 1.7) | 0 ( 0.0) | -0.017 | 5 ( 0.5) | 0 ( 0.0) | -0.005 | 46 ( 1.3) | 0 ( 0.0) | -0.013 |
| Fibrosis-4 Index (%) |  |  |  |  |  |  |  |  |  |
| < 1.45 | 550 (22.7) | 129 (21.7) | -0.010 | 162 (16.4) | 411 (14.7) | -0.017 | 712 (20.9) | 540 (15.9) | -0.049 |
| 1.45 – 3.25 | 994 (41.1) | 239 (40.2) | -0.009 | 394 (39.9) | 1260 (45.1) | 0.052 | 1388 (40.7) | 1499 (44.2) | 0.035 |
| 3.25 + | 697 (28.8) | 220 (37.0) | 0.082 | 377 (38.2) | 1081 (38.7) | 0.005 | 1074 (31.5) | 1301 (38.4) | 0.069 |
| Missing | 180 ( 7.4) | 7 ( 1.2) | -0.063 | 55 ( 5.6) | 41 ( 1.5) | -0.041 | 235 ( 6.9) | 48 ( 1.4) | -0.055 |
| Lactate, mmol/L (%) |  |  |  |  |  |  |  |  |  |
| 1_1.2 | 361 (14.9) | 118 (19.8) | 0.049 | 172 (17.4) | 496 (17.8) | 0.003 | 533 (15.6) | 614 (18.1) | 0.025 |
| 2_1.2LT2 | 514 (21.2) | 172 (28.9) | 0.077 | 254 (25.7) | 930 (33.3) | 0.076 | 768 (22.5) | 1102 (32.5) | 0.100 |
| 3_GE2 | 250 (10.3) | 96 (16.1) | 0.058 | 151 (15.3) | 413 (14.8) | -0.005 | 401 (11.8) | 509 (15.0) | 0.033 |
| Missing | 1296 (53.5) | 209 (35.1) | -0.184 | 411 (41.6) | 954 (34.2) | -0.074 | 1707 (50.1) | 1163 (34.3) | -0.157 |
| Platelet count per microL (%) |  |  |  |  |  |  |  |  |  |
| 150 or higher | 1631 (67.4) | 401 (67.4) | 0.000 | 613 (62.0) | 1893 (67.8) | 0.057 | 2244 (65.8) | 2294 (67.7) | 0.019 |
| < 150 | 744 (30.7) | 194 (32.6) | 0.019 | 371 (37.6) | 897 (32.1) | -0.054 | 1115 (32.7) | 1091 (32.2) | -0.005 |
| Missing | 46 ( 1.9) | 0 ( 0.0) | -0.019 | 4 ( 0.4) | 3 ( 0.1) | -0.003 | 50 ( 1.5) | 3 ( 0.1) | -0.014 |
| Total bilirubin, mg/dL (%) |  |  |  |  |  |  |  |  |  |
| < 1 | 1799 (74.3) | 456 (76.6) | 0.023 | 734 (74.3) | 2089 (74.8) | 0.005 | 2533 (74.3) | 2545 (75.1) | 0.008 |
| 1 - 1.2 | 182 ( 7.5) | 49 ( 8.2) | 0.007 | 94 ( 9.5) | 278 (10.0) | 0.004 | 276 ( 8.1) | 327 ( 9.7) | 0.016 |
| 1.2 + | 290 (12.0) | 86 (14.5) | 0.025 | 111 (11.2) | 393 (14.1) | 0.028 | 401 (11.8) | 479 (14.1) | 0.024 |
| Missing | 150 ( 6.2) | 4 ( 0.7) | -0.055 | 49 ( 5.0) | 33 ( 1.2) | -0.038 | 199 ( 5.8) | 37 ( 1.1) | -0.047 |
| White Blood Count per microL (%) |  |  |  |  |  |  |  |  |  |
| 4-10 | 1494 (61.7) | 274 (46.1) | -0.157 | 564 (57.1) | 1261 (45.1) | -0.119 | 2058 (60.4) | 1535 (45.3) | -0.151 |
| <4 | 606 (25.0) | 202 (33.9) | 0.089 | 269 (27.2) | 821 (29.4) | 0.022 | 875 (25.7) | 1023 (30.2) | 0.045 |
| >10 | 321 (13.3) | 119 (20.0) | 0.067 | 155 (15.7) | 711 (25.5) | 0.098 | 476 (14.0) | 830 (24.5) | 0.105 |
| C-reactive protein measured (%) | 1281 (52.9) | 415 (69.7) | 0.168 | 563 (57.0) | 1828 (65.4) | 0.085 | 1844 (54.1) | 2243 (66.2) | 0.121 |
| D-dimer measured (%) | 1712 (70.7) | 486 (81.7) | 0.110 | 775 (78.4) | 2354 (84.3) | 0.058 | 2487 (73.0) | 2840 (83.8) | 0.109 |
| **Viral Signs** |  |  |  |  |  |  |  |  |  |
| Highest Temperature (F) (%) |  |  |  |  |  |  |  |  |  |
| < 99 | 1051 (43.4) | 198 (33.3) | -0.101 | 279 (28.2) | 816 (29.2) | 0.010 | 1330 (39.0) | 1014 (29.9) | -0.091 |
| 99 - 100 | 606 (25.0) | 140 (23.5) | -0.015 | 230 (23.3) | 644 (23.1) | -0.002 | 836 (24.5) | 784 (23.1) | -0.014 |
| 100 - 102 | 537 (22.2) | 183 (30.8) | 0.086 | 319 (32.3) | 878 (31.4) | -0.009 | 856 (25.1) | 1061 (31.3) | 0.062 |
| 102 + | 220 ( 9.1) | 71 (11.9) | 0.028 | 158 (16.0) | 438 (15.7) | -0.003 | 378 (11.1) | 509 (15.0) | 0.039 |
| Missing | 7 ( 0.3) | 3 ( 0.5) | 0.002 | 2 ( 0.2) | 17 ( 0.6) | 0.004 | 9 ( 0.3) | 20 ( 0.6) | 0.003 |
| Mean Arterial Pressure, mmHg (%) |  |  |  |  |  |  |  |  |  |
| < 60 | 63 ( 2.6) | 9 ( 1.5) | -0.011 | 37 ( 3.7) | 46 ( 1.6) | -0.021 | 100 ( 2.9) | 55 ( 1.6) | -0.013 |
| 60 – 69 | 322 (13.3) | 51 ( 8.6) | -0.047 | 148 (15.0) | 341 (12.2) | -0.028 | 470 (13.8) | 392 (11.6) | -0.022 |
| 70 – 89 | 1575 (65.1) | 401 (67.4) | 0.023 | 643 (65.1) | 1906 (68.2) | 0.032 | 2218 (65.1) | 2307 (68.1) | 0.030 |
| 90 + | 455 (18.8) | 133 (22.4) | 0.036 | 159 (16.1) | 486 (17.4) | 0.013 | 614 (18.0) | 619 (18.3) | 0.003 |
| Missing | 6 ( 0.2) | 1 ( 0.2) | -0.001 | 1 ( 0.1) | 14 ( 0.5) | 0.004 | 7 ( 0.2) | 15 ( 0.4) | 0.002 |
| Lowest Oxygen Saturation (%) |  |  |  |  |  |  |  |  |  |
| < 88 | 30 ( 1.2) | 11 ( 1.8) | 0.006 | 78 ( 7.9) | 338 (12.1) | 0.042 | 108 ( 3.2) | 349 (10.3) | 0.071 |
| 88 - 92 | 630 (26.0) | 263 (44.2) | 0.182 | 489 (49.5) | 1686 (60.4) | 0.109 | 1119 (32.8) | 1949 (57.5) | 0.247 |
| 93 - 95 | 1198 (49.5) | 238 (40.0) | -0.095 | 311 (31.5) | 580 (20.8) | -0.107 | 1509 (44.3) | 818 (24.1) | -0.201 |
| 96 + | 508 (21.0) | 70 (11.8) | -0.092 | 92 ( 9.3) | 119 ( 4.3) | -0.051 | 600 (17.6) | 189 ( 5.6) | -0.120 |
| Missing | 55 ( 2.3) | 13 ( 2.2) | -0.001 | 18 ( 1.8) | 70 ( 2.5) | 0.007 | 73 ( 2.1) | 83 ( 2.4) | 0.003 |

**B. Propensity weighted pseudo population estimating the average treatment effect in the entire population (ATE)**

| Characteristics | No Oxygen | | | Nasal Canula | | | Combined Cohort | | |
| --- | --- | --- | --- | --- | --- | --- | --- | --- | --- |
|  | No | Yes | SMD | No | No | Yes | SMD | Yes | No |
| **Cohort, n** | 2979.1 | 2108.3 |  | 2585.7 | 3755.7 |  | 5796.7 | 6144.2 |  |
| **Age, (%)** |  |  |  |  |  |  |  |  |  |
| <50 | 324.5 (10.9) | 192.7 ( 9.1) | -0.018 | 205.9 ( 8.0) | 339.7 ( 9.0) | 0.011 | 514.4 ( 8.9) | 572.8 ( 9.3) | 0.004 |
| 50-59 | 368.8 (12.4) | 263.4 (12.5) | 0.001 | 314.0 (12.1) | 473.6 (12.6) | 0.005 | 683.7 (11.8) | 761.0 (12.4) | 0.006 |
| 60-69 | 686.9 (23.1) | 536.7 (25.5) | 0.024 | 569.7 (22.0) | 842.4 (22.4) | 0.004 | 1329.2 (22.9) | 1390.5 (22.6) | -0.003 |
| 70-79 | 964.0 (32.4) | 719.7 (34.1) | 0.018 | 938.2 (36.3) | 1384.5 (36.9) | 0.006 | 2055.2 (35.5) | 2247.7 (36.6) | 0.011 |
| 80+ | 634.9 (21.3) | 395.7 (18.8) | -0.025 | 557.9 (21.6) | 715.4 (19.0) | -0.025 | 1214.1 (20.9) | 1172.2 (19.1) | -0.019 |
| **Sex: Male, (%)** | 2811.8 (94.4) | 2014.8 (95.6) | 0.012 | 2438.8 (94.3) | 3558.1 (94.7) | 0.004 | 5458.7 (94.2) | 5808.7 (94.5) | 0.004 |
| **Race, (%)** |  |  |  |  |  |  |  |  |  |
| White, non-Hispanic | 1528.8 (51.3) | 1028.0 (48.8) | -0.026 | 1362.1 (52.7) | 2078.5 (55.3) | 0.027 | 3011.8 (52.0) | 3301.2 (53.7) | 0.018 |
| Black, non-Hispanic | 943.3 (31.7) | 686.9 (32.6) | 0.009 | 728.0 (28.2) | 984.7 (26.2) | -0.019 | 1759.4 (30.4) | 1731.6 (28.2) | -0.022 |
| Hispanic | 259.7 ( 8.7) | 212.2 (10.1) | 0.013 | 252.8 ( 9.8) | 355.2 ( 9.5) | -0.003 | 516.5 ( 8.9) | 577.9 ( 9.4) | 0.005 |
| Other | 149.9 ( 5.0) | 121.5 ( 5.8) | 0.007 | 159.1 ( 6.2) | 218.3 ( 5.8) | -0.003 | 317.7 ( 5.5) | 330.8 ( 5.4) | -0.001 |
| Unknown | 97.4 ( 3.3) | 59.7 ( 2.8) | -0.004 | 83.8 ( 3.2) | 119.0 ( 3.2) | -0.001 | 191.4 ( 3.3) | 202.9 ( 3.3) | 0.000 |
| **Phase (Admission Date) , (%)** |  |  |  |  |  |  |  |  |  |
| 1: June 7 - July 11 | 474.3 (15.9) | 273.5 (13.0) | -0.029 | 581.1 (22.5) | 602.7 (16.0) | -0.064 | 1077.2 (18.6) | 850.2 (13.8) | -0.047 |
| 2: July 12 - Aug. 15 | 515.4 (17.3) | 325.9 (15.5) | -0.018 | 538.5 (20.8) | 676.8 (18.0) | -0.028 | 1049.2 (18.1) | 1165.3 (19.0) | 0.009 |
| 3: Aug. 16 - Oct. 17 | 659.4 (22.1) | 483.1 (22.9) | 0.008 | 564.4 (21.8) | 772.6 (20.6) | -0.013 | 1260.0 (21.7) | 1290.7 (21.0) | -0.007 |
| 4: Oct. 18 - Dec. 5 | 1330.1 (44.6) | 1025.7 (48.6) | 0.040 | 901.7 (34.9) | 1703.6 (45.4) | 0.105 | 2410.3 (41.6) | 2838.0 (46.2) | 0.046 |
| **Site Dexamethasone Prescribing, (%)** |  |  |  |  |  |  |  |  |  |
| Low | 768.0 (25.8) | 365.1 (17.3) | -0.085 | 684.2 (26.5) | 719.1 (19.1) | -0.073 | 1477.9 (25.5) | 1227.7 (20.0) | -0.055 |
| Medium | 1811.7 (60.8) | 1379.7 (65.4) | 0.046 | 1510.0 (58.4) | 2236.0 (59.5) | 0.011 | 3565.1 (61.5) | 3705.7 (60.3) | -0.012 |
| High | 399.5 (13.4) | 363.5 (17.2) | 0.038 | 391.5 (15.1) | 800.5 (21.3) | 0.062 | 753.7 (13.0) | 1210.9 (19.7) | 0.067 |
| **Smoking Status, (%)** |  |  |  |  |  |  |  |  |  |
| Never Smoked | 1042.9 (35.0) | 761.9 (36.1) | 0.011 | 907.6 (35.1) | 1264.8 (33.7) | -0.014 | 2008.6 (34.7) | 2061.8 (33.6) | -0.011 |
| Former Smoker | 1149.4 (38.6) | 867.1 (41.1) | 0.025 | 1108.8 (42.9) | 1705.5 (45.4) | 0.025 | 2382.6 (41.1) | 2686.1 (43.7) | 0.026 |
| Current Smoker | 718.6 (24.1) | 452.5 (21.5) | -0.027 | 527.7 (20.4) | 721.5 (19.2) | -0.012 | 1315.7 (22.7) | 1275.6 (20.8) | -0.019 |
| Unknown | 68.3 ( 2.3) | 26.7 ( 1.3) | -0.010 | 41.6 ( 1.6) | 63.9 ( 1.7) | 0.001 | 89.8 ( 1.5) | 120.6 ( 2.0) | 0.004 |
| **AUDIT-C Score (%)** |  |  |  |  |  |  |  |  |  |
| 0 | 1836.1 (61.6) | 1303.7 (61.8) | 0.002 | 1690.4 (65.4) | 2422.7 (64.5) | -0.009 | 3677.0 (63.4) | 3870.0 (63.0) | -0.004 |
| 1 – 3 | 599.9 (20.1) | 435.8 (20.7) | 0.005 | 579.4 (22.4) | 872.6 (23.2) | 0.008 | 1236.9 (21.3) | 1393.5 (22.7) | 0.013 |
| 4 – 7 | 211.4 ( 7.1) | 161.7 ( 7.7) | 0.006 | 118.1 ( 4.6) | 180.5 ( 4.8) | 0.002 | 351.7 ( 6.1) | 348.8 ( 5.7) | -0.004 |
| 8 + | 92.9 ( 3.1) | 53.8 ( 2.6) | -0.006 | 44.7 ( 1.7) | 59.2 ( 1.6) | -0.002 | 130.9 ( 2.3) | 118.7 ( 1.9) | -0.003 |
| Unknown | 238.9 ( 8.0) | 153.3 ( 7.3) | -0.007 | 153.1 ( 5.9) | 220.8 ( 5.9) | 0.000 | 400.2 ( 6.9) | 413.2 ( 6.7) | -0.002 |
| **Comorbidities** |  |  |  |  |  |  |  |  |  |
| Myocardial Infarction (%) | 247.1 ( 8.3) | 138.3 ( 6.6) | -0.017 | 230.3 ( 8.9) | 310.2 ( 8.3) | -0.006 | 491.5 ( 8.5) | 468.9 ( 7.6) | -0.008 |
| Congestive Heart Failure (%) | 558.7 (18.8) | 411.2 (19.5) | 0.008 | 642.6 (24.9) | 806.2 (21.5) | -0.034 | 1257.0 (21.7) | 1256.7 (20.5) | -0.012 |
| Cerebrovascular Disease (%) | 534.4 (17.9) | 332.8 (15.8) | -0.022 | 451.3 (17.5) | 626.0 (16.7) | -0.008 | 1005.1 (17.3) | 988.3 (16.1) | -0.013 |
| Dementia (%) | 517.0 (17.4) | 270.8 (12.8) | -0.045 | 375.2 (14.5) | 439.1 (11.7) | -0.028 | 904.3 (15.6) | 721.8 (11.7) | -0.039 |
| Chronic Obstructive Pulmonary Disease (%) | 706.3 (23.7) | 570.9 (27.1) | 0.034 | 824.1 (31.9) | 1226.3 (32.7) | 0.008 | 1517.3 (26.2) | 1889.6 (30.8) | 0.046 |
| Rheumatoid Arthritis (%) | 50.9 ( 1.7) | 22.5 ( 1.1) | -0.006 | 54.0 ( 2.1) | 70.8 ( 1.9) | -0.002 | 79.5 ( 1.4) | 87.7 ( 1.4) | 0.001 |
| Peptic ulcer (%) | 78.9 ( 2.6) | 46.9 ( 2.2) | -0.004 | 50.8 ( 2.0) | 87.9 ( 2.3) | 0.004 | 133.3 ( 2.3) | 135.0 ( 2.2) | -0.001 |
| Liver disease, mild (%) | 397.6 (13.3) | 272.8 (12.9) | -0.004 | 239.1 ( 9.2) | 350.7 ( 9.3) | 0.001 | 637.9 (11.0) | 624.3 (10.2) | -0.008 |
| Diabetes, Uncomplicated (%) | 1327.3 (44.6) | 972.6 (46.1) | 0.016 | 1264.4 (48.9) | 1834.9 (48.9) | 0.000 | 2707.6 (46.7) | 2862.2 (46.6) | -0.001 |
| Diabetes, Complicated (%) | 862.3 (28.9) | 578.4 (27.4) | -0.015 | 837.4 (32.4) | 1128.6 (30.0) | -0.023 | 1810.1 (31.2) | 1773.0 (28.9) | -0.024 |
| Hemi or para plegia (%) | 94.4 ( 3.2) | 49.0 ( 2.3) | -0.008 | 74.6 ( 2.9) | 85.3 ( 2.3) | -0.006 | 168.6 ( 2.9) | 148.6 ( 2.4) | -0.005 |
| Liver disease, moderate-severe (%) | 73.8 ( 2.5) | 63.9 ( 3.0) | 0.006 | 27.1 ( 1.0) | 42.5 ( 1.1) | 0.001 | 95.4 ( 1.6) | 98.0 ( 1.6) | -0.001 |
| Metastatic cancer (%) | 62.3 ( 2.1) | 23.3 ( 1.1) | -0.010 | 32.3 ( 1.3) | 63.3 ( 1.7) | 0.004 | 121.9 ( 2.1) | 115.1 ( 1.9) | -0.002 |
| HIV (%) | 39.4 ( 1.3) | 29.1 ( 1.4) | 0.001 | 39.0 ( 1.5) | 51.2 ( 1.4) | -0.001 | 70.3 ( 1.2) | 57.5 ( 0.9) | -0.003 |
| Renal disease (%) | 735.1 (24.7) | 537.2 (25.5) | 0.008 | 729.4 (28.2) | 964.6 (25.7) | -0.025 | 1567.4 (27.0) | 1560.7 (25.4) | -0.016 |
| **Charlson Comorbidities Count (%)** |  |  |  |  |  |  |  |  |  |
| 0 | 621.0 (20.8) | 448.5 (21.3) | 0.004 | 485.5 (18.8) | 689.1 (18.3) | -0.004 | 1122.3 (19.4) | 1207.4 (19.7) | 0.003 |
| 1 - 2 | 920.5 (30.9) | 688.0 (32.6) | 0.017 | 733.7 (28.4) | 1184.0 (31.5) | 0.032 | 1767.9 (30.5) | 1953.9 (31.8) | 0.013 |
| 3 - 4 | 654.1 (22.0) | 456.2 (21.6) | -0.003 | 617.4 (23.9) | 913.0 (24.3) | 0.004 | 1327.6 (22.9) | 1481.3 (24.1) | 0.012 |
| 5 + | 783.5 (26.3) | 515.6 (24.5) | -0.018 | 749.1 (29.0) | 969.6 (25.8) | -0.032 | 1578.9 (27.2) | 1501.7 (24.4) | -0.028 |
| **Number of Doctors (prior year) (%)** |  |  |  |  |  |  |  |  |  |
| 0 | 1276.8 (42.9) | 885.2 (42.0) | -0.009 | 985.6 (38.1) | 1430.6 (38.1) | 0.000 | 2330.1 (40.2) | 2475.4 (40.3) | 0.001 |
| 1 | 816.9 (27.4) | 627.3 (29.8) | 0.023 | 674.8 (26.1) | 1043.7 (27.8) | 0.017 | 1588.7 (27.4) | 1685.4 (27.4) | 0.000 |
| 2 - 4 | 812.5 (27.3) | 555.7 (26.4) | -0.009 | 850.5 (32.9) | 1196.5 (31.9) | -0.010 | 1717.9 (29.6) | 1835.7 (29.9) | 0.002 |
| 5 + | 72.9 ( 2.4) | 40.1 ( 1.9) | -0.005 | 74.8 ( 2.9) | 84.9 ( 2.3) | -0.006 | 160.0 ( 2.8) | 147.6 ( 2.4) | -0.004 |
| **Specialty clinics attended**  Cardiology **(%)** | 732.6 (24.6) | 531.3 (25.2) | 0.006 | 741.9 (28.7) | 1059.3 (28.2) | -0.005 | 1512.0 (26.1) | 1661.1 (27.0) | 0.010 |
| Coagulation (%) | 47.5 ( 1.6) | 46.8 ( 2.2) | 0.006 | 40.3 ( 1.6) | 49.8 ( 1.3) | -0.002 | 88.0 ( 1.5) | 93.8 ( 1.5) | 0.000 |
| Pacemaker (%) | 112.6 ( 3.8) | 115.5 ( 5.5) | 0.017 | 92.3 ( 3.6) | 115.7 ( 3.1) | -0.005 | 232.5 ( 4.0) | 220.8 ( 3.6) | -0.004 |
| Dialysis (%) | 34.9 ( 1.2) | 21.2 ( 1.0) | -0.002 | 70.4 ( 2.7) | 66.6 ( 1.8) | -0.010 | 98.7 ( 1.7) | 99.8 ( 1.6) | -0.001 |
| Gastoenterology (%) | 256.1 ( 8.6) | 175.1 ( 8.3) | -0.003 | 234.8 ( 9.1) | 368.2 ( 9.8) | 0.007 | 508.1 ( 8.8) | 586.5 ( 9.5) | 0.008 |
| Hepatology (%) | 105.7 ( 3.5) | 67.7 ( 3.2) | -0.003 | 77.4 ( 3.0) | 94.7 ( 2.5) | -0.005 | 183.2 ( 3.2) | 146.2 ( 2.4) | -0.008 |
| Homeless (%) | 231.8 ( 7.8) | 148.3 ( 7.0) | -0.007 | 127.0 ( 4.9) | 141.8 ( 3.8) | -0.011 | 376.3 ( 6.5) | 304.9 ( 5.0) | -0.015 |
| **Co-medications** |  |  |  |  |  |  |  |  |  |
| Prophylactic Anticoagulants | 1597.7 (53.6) | 1127.2 (53.5) | -0.002 | 1488.4 (57.6) | 2171.8 (57.8) | 0.003 | 3164.0 (54.6) | 3444.3 (56.1) | 0.015 |
| Remdesivir, 1^st^ 48 hours (%) | 433.7 (14.6) | 439.7 (20.9) | 0.063 | 999.8 (38.7) | 2174.3 (57.9) | 0.192 | 1714.5 (29.6) | 2630.2 (42.8) | 0.132 |
| **Laboratory Results** |  |  |  |  |  |  |  |  |  |
| Albumin, g/dL (%) |  |  |  |  |  |  |  |  |  |
| 3.5 + | 1267.1 (42.5) | 770.3 (36.5) | -0.060 | 795.3 (30.8) | 1084.5 (28.9) | -0.019 | 2125.2 (36.7) | 2040.4 (33.2) | -0.035 |
| 3 - 3.49 | 924.4 (31.0) | 728.9 (34.6) | 0.035 | 878.8 (34.0) | 1348.7 (35.9) | 0.019 | 1871.1 (32.3) | 2103.8 (34.2) | 0.020 |
| < 3 | 622.6 (20.9) | 529.5 (25.1) | 0.042 | 811.5 (31.4) | 1199.6 (31.9) | 0.006 | 1527.9 (26.4) | 1775.3 (28.9) | 0.025 |
| Missing | 165.0 ( 5.5) | 79.6 ( 3.8) | -0.018 | 100.1 ( 3.9) | 122.9 ( 3.3) | -0.006 | 272.5 ( 4.7) | 224.8 ( 3.7) | -0.010 |
| Alanine aminotransferase, IU/L (%) |  |  |  |  |  |  |  |  |  |
| < 20 | 949.8 (31.9) | 596.0 (28.3) | -0.036 | 656.3 (25.4) | 853.0 (22.7) | -0.027 | 1678.7 (29.0) | 1532.1 (24.9) | -0.040 |
| 20 - 39 | 1155.8 (38.8) | 865.1 (41.0) | 0.022 | 1137.9 (44.0) | 1638.7 (43.6) | -0.004 | 2415.7 (41.7) | 2621.7 (42.7) | 0.010 |
| 40 + | 731.2 (24.5) | 613.5 (29.1) | 0.046 | 714.2 (27.6) | 1184.6 (31.5) | 0.039 | 1494.2 (25.8) | 1839.4 (29.9) | 0.042 |
| Missing | 142.3 ( 4.8) | 33.6 ( 1.6) | -0.032 | 77.2 ( 3.0) | 79.4 ( 2.1) | -0.009 | 208.1 ( 3.6) | 151.1 ( 2.5) | -0.011 |
| Asparate aminostransferase, IU/L (%) |  |  |  |  |  |  |  |  |  |
| < 20 | 699.1 (23.5) | 272.4 (12.9) | -0.105 | 331.1 (12.8) | 399.0 (10.6) | -0.022 | 1054.7 (18.2) | 829.8 (13.5) | -0.047 |
| 20 - 39 | 1372.6 (46.1) | 1075.9 (51.0) | 0.050 | 1166.3 (45.1) | 1679.2 (44.7) | -0.004 | 2670.2 (46.1) | 2816.1 (45.8) | -0.002 |
| 40 + | 907.4 (30.5) | 760.0 (36.0) | 0.056 | 1088.4 (42.1) | 1677.6 (44.7) | 0.026 | 2071.8 (35.7) | 2498.3 (40.7) | 0.049 |
| Creatinine, mg/dL (%) |  |  |  |  |  |  |  |  |  |
| < 1.2 | 1499.4 (50.3) | 967.2 (45.9) | -0.045 | 1108.0 (42.9) | 1704.5 (45.4) | 0.025 | 2671.7 (46.1) | 2857.0 (46.5) | 0.004 |
| 1.2 – 1.99 | 1035.8 (34.8) | 811.3 (38.5) | 0.037 | 992.1 (38.4) | 1432.2 (38.1) | -0.002 | 2141.5 (36.9) | 2265.1 (36.9) | -0.001 |
| 2 + | 432.9 (14.5) | 329.8 (15.6) | 0.011 | 485.6 (18.8) | 619.0 (16.5) | -0.023 | 983.5 (17.0) | 1022.1 (16.6) | -0.003 |
| Missing | 11.0 ( 0.4) | 0.0 ( 0.0) | -0.004 | 0.0 ( 0.0) | 0.0 ( 0.0) |  | 0.0 ( 0.0) | 0.0 ( 0.0) |  |
| Fibrosis-4 Index (%) |  |  |  |  |  |  |  |  |  |
| < 1.45 | 686.8 (23.1) | 439.5 (20.8) | -0.022 | 353.2 (13.7) | 569.2 (15.2) | 0.015 | 1065.6 (18.4) | 1062.8 (17.3) | -0.011 |
| 1.45 – 3.25 | 1204.6 (40.4) | 837.1 (39.7) | -0.007 | 1067.9 (41.3) | 1644.5 (43.8) | 0.025 | 2393.0 (41.3) | 2597.0 (42.3) | 0.010 |
| 3.25 + | 928.5 (31.2) | 798.1 (37.9) | 0.067 | 1083.7 (41.9) | 1456.9 (38.8) | -0.031 | 2101.8 (36.3) | 2325.3 (37.8) | 0.016 |
| Missing | 159.2 ( 5.3) | 33.6 ( 1.6) | -0.038 | 81.0 ( 3.1) | 85.1 ( 2.3) | -0.009 | 236.3 ( 4.1) | 159.1 ( 2.6) | -0.015 |
| Lactate, mmol/L (%) |  |  |  |  |  |  |  |  |  |
| 1_1.2 | 493.0 (16.5) | 441.7 (21.0) | 0.044 | 455.4 (17.6) | 695.1 (18.5) | 0.009 | 1003.8 (17.3) | 1075.5 (17.5) | 0.002 |
| 2_1.2LT2 | 682.6 (22.9) | 558.3 (26.5) | 0.036 | 713.6 (27.6) | 1170.2 (31.2) | 0.036 | 1510.3 (26.1) | 1816.4 (29.6) | 0.035 |
| 3_GE2 | 348.1 (11.7) | 312.3 (14.8) | 0.031 | 385.0 (14.9) | 559.1 (14.9) | 0.000 | 737.4 (12.7) | 880.6 (14.3) | 0.016 |
| Missing | 1455.5 (48.9) | 796.0 (37.8) | -0.111 | 1031.7 (39.9) | 1331.3 (35.4) | -0.045 | 2545.2 (43.9) | 2371.8 (38.6) | -0.053 |
| Platelet count per microL (%) |  |  |  |  |  |  |  |  |  |
| 150 or higher | 2019.9 (67.8) | 1370.8 (65.0) | -0.028 | 1636.0 (63.3) | 2488.6 (66.3) | 0.030 | 3822.5 (65.9) | 4050.8 (65.9) | 0.000 |
| < 150 | 943.2 (31.7) | 737.4 (35.0) | 0.033 | 946.9 (36.6) | 1262.6 (33.6) | -0.030 | 1964.6 (33.9) | 2081.7 (33.9) | 0.000 |
| Missing | 16.0 ( 0.5) | 0.0 ( 0.0) | -0.005 | 2.9 ( 0.1) | 4.5 ( 0.1) | 0.000 | 9.6 ( 0.2) | 11.7 ( 0.2) | 0.000 |
| Total bilirubin, mg/dL (%) |  |  |  |  |  |  |  |  |  |
| < 1 | 2268.0 (76.1) | 1624.2 (77.0) | 0.009 | 1978.0 (76.5) | 2841.7 (75.7) | -0.008 | 4395.1 (75.8) | 4646.6 (75.6) | -0.002 |
| 1 - 1.2 | 218.7 ( 7.3) | 180.6 ( 8.6) | 0.012 | 228.2 ( 8.8) | 354.7 ( 9.4) | 0.006 | 489.1 ( 8.4) | 558.8 ( 9.1) | 0.007 |
| 1.2 + | 368.0 (12.4) | 281.1 (13.3) | 0.010 | 307.6 (11.9) | 487.2 (13.0) | 0.011 | 730.4 (12.6) | 808.8 (13.2) | 0.006 |
| Missing | 124.4 ( 4.2) | 22.3 ( 1.1) | -0.031 | 71.9 ( 2.8) | 72.2 ( 1.9) | -0.009 | 182.1 ( 3.1) | 130.1 ( 2.1) | -0.010 |
| White Blood Count per microL (%) |  |  |  |  |  |  |  |  |  |
| 4-10 | 1746.6 (58.6) | 1082.6 (51.3) | -0.073 | 1378.3 (53.3) | 1818.5 (48.4) | -0.049 | 3243.2 (55.9) | 3097.6 (50.4) | -0.055 |
| <4 | 803.4 (27.0) | 653.7 (31.0) | 0.040 | 769.2 (29.7) | 1095.3 (29.2) | -0.006 | 1622.3 (28.0) | 1776.7 (28.9) | 0.009 |
| >10 | 429.1 (14.4) | 372.0 (17.6) | 0.032 | 438.2 (16.9) | 841.9 (22.4) | 0.055 | 931.2 (16.1) | 1269.8 (20.7) | 0.046 |
| C-reactive protein measured (%) | 1689.4 (56.7) | 1364.5 (64.7) | 0.080 | 1550.1 (59.9) | 2386.3 (63.5) | 0.036 | 3428.1 (59.1) | 3883.6 (63.2) | 0.041 |
| D-dimer measured (%) | 2184.7 (73.3) | 1657.5 (78.6) | 0.053 | 2106.3 (81.5) | 3153.8 (84.0) | 0.025 | 4512.3 (77.8) | 4998.2 (81.3) | 0.035 |
| **Vital Signs** |  |  |  |  |  |  |  |  |  |
| Highest Temperature (F) (%) |  |  |  |  |  |  |  |  |  |
| < 99 | 1226.6 (41.2) | 832.8 (39.5) | -0.017 | 684.9 (26.5) | 1071.6 (28.5) | 0.020 | 1968.5 (34.0) | 2001.3 (32.6) | -0.014 |
| 99 - 100 | 735.0 (24.7) | 479.8 (22.8) | -0.019 | 542.6 (21.0) | 846.3 (22.5) | 0.015 | 1389.2 (24.0) | 1452.8 (23.6) | -0.003 |
| 100 - 102 | 693.4 (23.3) | 522.8 (24.8) | 0.015 | 869.7 (33.6) | 1231.0 (32.8) | -0.009 | 1601.0 (27.6) | 1769.6 (28.8) | 0.012 |
| 102 + | 314.2 (10.5) | 267.2 (12.7) | 0.021 | 478.5 (18.5) | 591.4 (15.7) | -0.028 | 818.1 (14.1) | 893.9 (14.5) | 0.004 |
| Missing | 9.8 ( 0.3) | 5.8 ( 0.3) | -0.001 | 9.9 ( 0.4) | 15.4 ( 0.4) | 0.000 | 19.9 ( 0.3) | 26.7 ( 0.4) | 0.001 |
| Mean Arterial Pressure, mmHg (%) |  |  |  |  |  |  |  |  |  |
| < 60 | 70.8 ( 2.4) | 53.4 ( 2.5) | 0.002 | 61.1 ( 2.4) | 72.9 ( 1.9) | -0.004 | 141.0 ( 2.4) | 118.5 ( 1.9) | -0.005 |
| 60 – 69 | 365.0 (12.3) | 205.2 ( 9.7) | -0.025 | 387.6 (15.0) | 509.2 (13.6) | -0.014 | 784.4 (13.5) | 751.0 (12.2) | -0.013 |
| 70 – 89 | 1954.5 (65.6) | 1416.8 (67.2) | 0.016 | 1721.5 (66.6) | 2531.4 (67.4) | 0.008 | 3812.6 (65.8) | 4141.8 (67.4) | 0.016 |
| 90 + | 582.2 (19.5) | 431.2 (20.5) | 0.009 | 407.1 (15.7) | 630.2 (16.8) | 0.010 | 1049.4 (18.1) | 1112.1 (18.1) | 0.000 |
| Missing | 6.6 ( 0.2) | 1.7 ( 0.1) | -0.001 | 8.5 ( 0.3) | 12.1 ( 0.3) | 0.000 | 9.3 ( 0.2) | 20.8 ( 0.3) | 0.002 |
| Lowest Oxygen Saturation (%) |  |  |  |  |  |  |  |  |  |
| < 88 | 43.0 ( 1.4) | 25.2 ( 1.2) | -0.002 | 245.8 ( 9.5) | 415.6 (11.1) | 0.016 | 312.7 ( 5.4) | 468.1 ( 7.6) | 0.022 |
| 88 - 92 | 887.9 (29.8) | 707.3 (33.6) | 0.037 | 1410.9 (54.6) | 2170.9 (57.8) | 0.032 | 2438.6 (42.1) | 3022.9 (49.2) | 0.071 |
| 93 - 95 | 1415.5 (47.5) | 997.6 (47.3) | -0.002 | 727.3 (28.1) | 896.6 (23.9) | -0.043 | 2191.5 (37.8) | 2006.9 (32.7) | -0.051 |
| 96 + | 564.1 (18.9) | 307.0 (14.6) | -0.044 | 156.6 ( 6.1) | 193.7 ( 5.2) | -0.009 | 733.5 (12.7) | 513.5 ( 8.4) | -0.043 |
| Missing | 68.6 ( 2.3) | 71.2 ( 3.4) | 0.011 | 45.1 ( 1.7) | 78.9 ( 2.1) | 0.004 | 120.3 ( 2.1) | 132.9 ( 2.2) | 0.001 |

**C. Propensity weighted pseudo population estimating the average treatment effect in the treated population (ATT)**

| Characteristics | No Oxygen | | | Nasal Canula | | | Combined Cohort | | |
| --- | --- | --- | --- | --- | --- | --- | --- | --- | --- |
|  | No | Yes | SMD | No | No | Yes | SMD | Yes | No |
| **Cohort, n** | 452.7 | 595.0 |  | 1638.9 | 2793.0 |  | 2200.7 | 3388.0 |  |
| **Age, (%)** |  |  |  |  |  |  |  |  |  |
| <50 | 39.7 ( 8.8) | 59.0 ( 9.9) | 0.011 | 138.8 ( 8.5) | 261.0 ( 9.3) | 0.009 | 177.4 ( 8.1) | 320.0 ( 9.4) | 0.014 |
| 50-59 | 60.0 (13.3) | 90.0 (15.1) | 0.019 | 192.7 (11.8) | 344.0 (12.3) | 0.006 | 262.4 (11.9) | 434.0 (12.8) | 0.009 |
| 60-69 | 113.7 (25.1) | 138.0 (23.2) | -0.019 | 360.3 (22.0) | 647.0 (23.2) | 0.012 | 496.8 (22.6) | 785.0 (23.2) | 0.006 |
| 70-79 | 150.3 (33.2) | 195.0 (32.8) | -0.004 | 611.7 (37.3) | 1048.0 (37.5) | 0.002 | 802.7 (36.5) | 1243.0 (36.7) | 0.002 |
| 80+ | 88.9 (19.6) | 113.0 (19.0) | -0.007 | 335.5 (20.5) | 493.0 (17.7) | -0.028 | 461.4 (21.0) | 606.0 (17.9) | -0.031 |
| **Sex: Male, (%)** | 430.0 (95.0) | 562.0 (94.5) | -0.005 | 1545.1 (94.3) | 2644.0 (94.7) | 0.004 | 2069.5 (94.0) | 3206.0 (94.6) | 0.006 |
| **Race, (%)** |  |  |  |  |  |  |  |  |  |
| White, non-Hispanic | 241.0 (53.2) | 321.0 (53.9) | 0.007 | 865.2 (52.8) | 1589.0 (56.9) | 0.041 | 1174.2 (53.4) | 1910.0 (56.4) | 0.030 |
| Black, non-Hispanic | 134.0 (29.6) | 159.0 (26.7) | -0.029 | 438.4 (26.7) | 678.0 (24.3) | -0.025 | 612.6 (27.8) | 837.0 (24.7) | -0.031 |
| Hispanic | 41.5 ( 9.2) | 61.0 (10.3) | 0.011 | 175.6 (10.7) | 290.0 (10.4) | -0.003 | 209.1 ( 9.5) | 351.0 (10.4) | 0.009 |
| Other | 22.8 ( 5.0) | 28.0 ( 4.7) | -0.003 | 104.0 ( 6.3) | 152.0 ( 5.4) | -0.009 | 140.7 ( 6.4) | 180.0 ( 5.3) | -0.011 |
| Unknown | 13.3 ( 2.9) | 26.0 ( 4.4) | 0.014 | 55.8 ( 3.4) | 84.0 ( 3.0) | -0.004 | 64.1 ( 2.9) | 110.0 ( 3.2) | 0.003 |
| **Phase (Admission Date) , (%)** |  |  |  |  |  |  |  |  |  |
| 1: June 7 - July 11 | 52.3 (11.6) | 51.0 ( 8.6) | -0.030 | 294.7 (18.0) | 270.0 ( 9.7) | -0.083 | 361.2 (16.4) | 321.0 ( 9.5) | -0.069 |
| 2: July 12 - Aug. 15 | 63.6 (14.1) | 79.0 (13.3) | -0.008 | 343.2 (20.9) | 490.0 (17.5) | -0.034 | 380.2 (17.3) | 569.0 (16.8) | -0.005 |
| 3: Aug. 16 - Oct. 17 | 100.9 (22.3) | 139.0 (23.4) | 0.011 | 368.3 (22.5) | 587.0 (21.0) | -0.015 | 457.7 (20.8) | 726.0 (21.4) | 0.006 |
| 4: Oct. 18 - Dec. 5 | 235.9 (52.1) | 326.0 (54.8) | 0.027 | 632.7 (38.6) | 1446.0 (51.8) | 0.132 | 1001.6 (45.5) | 1772.0 (52.3) | 0.068 |
| **Site Dexamethasone Prescribing, (%)** |  |  |  |  |  |  |  |  |  |
| Low | 79.3 (17.5) | 84.0 (14.1) | -0.034 | 392.3 (23.9) | 421.0 (15.1) | -0.089 | 481.9 (21.9) | 505.0 (14.9) | -0.070 |
| Medium | 290.3 (64.1) | 358.0 (60.2) | -0.040 | 964.2 (58.8) | 1665.0 (59.6) | 0.008 | 1353.7 (61.5) | 2023.0 (59.7) | -0.018 |
| High | 83.0 (18.3) | 153.0 (25.7) | 0.074 | 282.4 (17.2) | 707.0 (25.3) | 0.081 | 365.1 (16.6) | 860.0 (25.4) | 0.088 |
| **Smoking Status, (%)** |  |  |  |  |  |  |  |  |  |
| Never Smoked | 157.4 (34.8) | 203.0 (34.1) | -0.007 | 581.8 (35.5) | 939.0 (33.6) | -0.019 | 782.8 (35.6) | 1142.0 (33.7) | -0.019 |
| Former Smoker | 188.9 (41.7) | 269.0 (45.2) | 0.035 | 720.5 (44.0) | 1305.0 (46.7) | 0.028 | 979.9 (44.5) | 1574.0 (46.5) | 0.019 |
| Current Smoker | 99.6 (22.0) | 112.0 (18.8) | -0.032 | 323.2 (19.7) | 506.0 (18.1) | -0.016 | 415.3 (18.9) | 618.0 (18.2) | -0.006 |
| Unknown | 6.8 ( 1.5) | 11.0 ( 1.8) | 0.004 | 13.5 ( 0.8) | 43.0 ( 1.5) | 0.007 | 22.8 ( 1.0) | 54.0 ( 1.6) | 0.006 |
| **AUDIT-C Score (%)** |  |  |  |  |  |  |  |  |  |
| 0 | 280.2 (61.9) | 369.0 (62.0) | 0.001 | 1068.2 (65.2) | 1767.0 (63.3) | -0.019 | 1413.4 (64.2) | 2136.0 (63.0) | -0.012 |
| 1 – 3 | 94.0 (20.8) | 132.0 (22.2) | 0.014 | 389.1 (23.7) | 674.0 (24.1) | 0.004 | 501.8 (22.8) | 806.0 (23.8) | 0.010 |
| 4 – 7 | 29.7 ( 6.6) | 34.0 ( 5.7) | -0.009 | 82.1 ( 5.0) | 147.0 ( 5.3) | 0.003 | 132.4 ( 6.0) | 181.0 ( 5.3) | -0.007 |
| 8 + | 11.9 ( 2.6) | 15.0 ( 2.5) | -0.001 | 24.6 ( 1.5) | 43.0 ( 1.5) | 0.000 | 29.9 ( 1.4) | 58.0 ( 1.7) | 0.004 |
| Unknown | 36.9 ( 8.1) | 45.0 ( 7.6) | -0.006 | 74.9 ( 4.6) | 162.0 ( 5.8) | 0.012 | 123.2 ( 5.6) | 207.0 ( 6.1) | 0.005 |
| **Comorbidities** |  |  |  |  |  |  |  |  |  |
| Myocardial Infarction (%) | 30.4 ( 6.7) | 36.0 ( 6.1) | -0.007 | 144.2 ( 8.8) | 216.0 ( 7.7) | -0.011 | 183.4 ( 8.3) | 252.0 ( 7.4) | -0.009 |
| Congestive Heart Failure (%) | 83.8 (18.5) | 102.0 (17.1) | -0.014 | 381.8 (23.3) | 534.0 (19.1) | -0.042 | 495.4 (22.5) | 636.0 (18.8) | -0.037 |
| Cerebrovascular Disease (%) | 66.4 (14.7) | 79.0 (13.3) | -0.014 | 271.0 (16.5) | 418.0 (15.0) | -0.016 | 344.9 (15.7) | 497.0 (14.7) | -0.010 |
| Dementia (%) | 47.8 (10.6) | 59.0 ( 9.9) | -0.007 | 207.7 (12.7) | 252.0 ( 9.0) | -0.036 | 268.3 (12.2) | 311.0 ( 9.2) | -0.030 |
| Chronic Obstructive Pulmonary Disease (%) | 118.9 (26.3) | 168.0 (28.2) | 0.020 | 530.1 (32.3) | 923.0 (33.0) | 0.007 | 615.9 (28.0) | 1091.0 (32.2) | 0.042 |
| Rheumatoid Arthritis (%) | 5.8 ( 1.3) | 9.0 ( 1.5) | 0.002 | 40.0 ( 2.4) | 49.0 ( 1.8) | -0.007 | 34.5 ( 1.6) | 58.0 ( 1.7) | 0.001 |
| Peptic ulcer (%) | 7.9 ( 1.7) | 9.0 ( 1.5) | -0.002 | 27.8 ( 1.7) | 55.0 ( 2.0) | 0.003 | 39.2 ( 1.8) | 64.0 ( 1.9) | 0.001 |
| Liver disease, mild (%) | 53.6 (11.8) | 66.0 (11.1) | -0.008 | 141.9 ( 8.7) | 265.0 ( 9.5) | 0.008 | 206.1 ( 9.4) | 331.0 ( 9.8) | 0.004 |
| Diabetes, Uncomplicated (%) | 207.8 (45.9) | 278.0 (46.7) | 0.008 | 806.5 (49.2) | 1369.0 (49.0) | -0.002 | 1072.6 (48.7) | 1647.0 (48.6) | -0.001 |
| Diabetes, Complicated (%) | 132.1 (29.2) | 168.0 (28.2) | -0.010 | 530.6 (32.4) | 819.0 (29.3) | -0.031 | 679.9 (30.9) | 987.0 (29.1) | -0.018 |
| Hemi or paraplegia (%) | 11.4 ( 2.5) | 10.0 ( 1.7) | -0.008 | 38.6 ( 2.4) | 52.0 ( 1.9) | -0.005 | 50.5 ( 2.3) | 62.0 ( 1.8) | -0.005 |
| Liver disease, moderate-severe (%) | 8.6 ( 1.9) | 9.0 ( 1.5) | -0.004 | 14.1 ( 0.9) | 32.0 ( 1.1) | 0.003 | 28.4 ( 1.3) | 41.0 ( 1.2) | -0.001 |
| Metastatic cancer (%) | 6.3 ( 1.4) | 6.0 ( 1.0) | -0.004 | 18.3 ( 1.1) | 46.0 ( 1.6) | 0.005 | 49.9 ( 2.3) | 52.0 ( 1.5) | -0.007 |
| HIV (%) | 7.4 ( 1.6) | 10.0 ( 1.7) | 0.000 | 26.9 ( 1.6) | 20.0 ( 0.7) | -0.009 | 27.3 ( 1.2) | 30.0 ( 0.9) | -0.004 |
| Renal disease (%) | 113.0 (25.0) | 140.0 (23.5) | -0.014 | 456.8 (27.9) | 656.0 (23.5) | -0.044 | 599.0 (27.2) | 796.0 (23.5) | -0.037 |
| **Charlson Comorbidities Count (%)** |  |  |  |  |  |  |  |  |  |
| 0 | 92.5 (20.4) | 134.0 (22.5) | 0.021 | 314.0 (19.2) | 518.0 (18.5) | -0.006 | 426.0 (19.4) | 652.0 (19.2) | -0.001 |
| 1 - 2 | 150.2 (33.2) | 200.0 (33.6) | 0.004 | 469.4 (28.6) | 934.0 (33.4) | 0.048 | 692.5 (31.5) | 1134.0 (33.5) | 0.020 |
| 3 - 4 | 106.0 (23.4) | 131.0 (22.0) | -0.014 | 392.1 (23.9) | 702.0 (25.1) | 0.012 | 484.5 (22.0) | 833.0 (24.6) | 0.026 |
| 5 + | 104.1 (23.0) | 130.0 (21.8) | -0.011 | 463.4 (28.3) | 639.0 (22.9) | -0.054 | 597.7 (27.2) | 769.0 (22.7) | -0.045 |
| **Number of Doctors (prior year) (%)** |  |  |  |  |  |  |  |  |  |
| 0 | 185.8 (41.1) | 250.0 (42.0) | 0.010 | 604.6 (36.9) | 1075.0 (38.5) | 0.016 | 842.3 (38.3) | 1325.0 (39.1) | 0.008 |
| 1 | 138.4 (30.6) | 182.0 (30.6) | 0.000 | 440.5 (26.9) | 780.0 (27.9) | 0.010 | 610.1 (27.7) | 962.0 (28.4) | 0.007 |
| 2 - 4 | 121.6 (26.9) | 155.0 (26.1) | -0.008 | 549.1 (33.5) | 879.0 (31.5) | -0.020 | 683.3 (31.0) | 1034.0 (30.5) | -0.005 |
| 5 + | 6.9 ( 1.5) | 8.0 ( 1.3) | -0.002 | 44.7 ( 2.7) | 59.0 ( 2.1) | -0.006 | 65.0 ( 3.0) | 67.0 ( 2.0) | -0.010 |
| **Specialty clinics attended**  Cardiology **(%)** | 109.1 (24.1) | 147.0 (24.7) | 0.006 | 461.5 (28.2) | 759.0 (27.2) | -0.010 | 599.5 (27.2) | 906.0 (26.7) | -0.005 |
| Coagulation (%) | 9.5 ( 2.1) | 13.0 ( 2.2) | 0.001 | 20.2 ( 1.2) | 31.0 ( 1.1) | -0.001 | 30.0 ( 1.4) | 44.0 ( 1.3) | -0.001 |
| Pacemaker (%) | 15.2 ( 3.4) | 20.0 ( 3.4) | 0.000 | 45.2 ( 2.8) | 70.0 ( 2.5) | -0.002 | 81.6 ( 3.7) | 90.0 ( 2.7) | -0.011 |
| Dialysis (%) | 3.9 ( 0.9) | 5.0 ( 0.8) | 0.000 | 38.2 ( 2.3) | 33.0 ( 1.2) | -0.012 | 35.7 ( 1.6) | 38.0 ( 1.1) | -0.005 |
| Gastoenterology (%) | 41.7 ( 9.2) | 61.0 (10.3) | 0.010 | 150.7 ( 9.2) | 292.0 (10.5) | 0.013 | 216.0 ( 9.8) | 353.0 (10.4) | 0.006 |
| Hepatology (%) | 9.7 ( 2.2) | 10.0 ( 1.7) | -0.005 | 47.3 ( 2.9) | 64.0 ( 2.3) | -0.006 | 59.2 ( 2.7) | 74.0 ( 2.2) | -0.005 |
| Homeless (%) | 27.1 ( 6.0) | 30.0 ( 5.0) | -0.009 | 70.7 ( 4.3) | 93.0 ( 3.3) | -0.010 | 112.4 ( 5.1) | 123.0 ( 3.6) | -0.015 |
| **Co-medications** |  |  |  |  |  |  |  |  |  |
| Prophylactic Anticoagulants | 248.2 (54.8) | 332.0 (55.8) | 0.010 | 983.7 (60.0) | 1643.0 (58.8) | -0.012 | 1231.8 (56.0) | 1975.0 (58.3) | 0.023 |
| Remdesivir, 1^st^ 48 hours (%) | 124.3 (27.5) | 257.0 (43.2) | 0.157 | 826.8 (50.4) | 2001.0 (71.6) | 0.212 | 1087.6 (49.4) | 2258.0 (66.6) | 0.172 |
| **Laboratory Results** |  |  |  |  |  |  |  |  |  |
| Albumin, g/dL (%) |  |  |  |  |  |  |  |  |  |
| 3.5 + | 174.5 (38.6) | 218.0 (36.6) | -0.019 | 498.7 (30.4) | 781.0 (28.0) | -0.025 | 677.5 (30.8) | 999.0 (29.5) | -0.013 |
| 3 - 3.49 | 151.9 (33.5) | 203.0 (34.1) | 0.006 | 551.1 (33.6) | 1005.0 (36.0) | 0.024 | 773.8 (35.2) | 1208.0 (35.7) | 0.005 |
| < 3 | 107.9 (23.8) | 151.0 (25.4) | 0.015 | 537.1 (32.8) | 928.0 (33.2) | 0.005 | 680.4 (30.9) | 1079.0 (31.8) | 0.009 |
| Missing | 18.4 ( 4.1) | 23.0 ( 3.9) | -0.002 | 51.9 ( 3.2) | 79.0 ( 2.8) | -0.003 | 69.0 ( 3.1) | 102.0 ( 3.0) | -0.001 |
| Alanine aminotransferase, IU/L (%) |  |  |  |  |  |  |  |  |  |
| < 20 | 129.5 (28.6) | 148.0 (24.9) | -0.037 | 370.4 (22.6) | 578.0 (20.7) | -0.019 | 540.9 (24.6) | 726.0 (21.4) | -0.031 |
| 20 - 39 | 187.3 (41.4) | 242.0 (40.7) | -0.007 | 737.3 (45.0) | 1201.0 (43.0) | -0.020 | 936.1 (42.5) | 1443.0 (42.6) | 0.001 |
| 40 + | 129.6 (28.6) | 199.0 (33.4) | 0.048 | 502.2 (30.6) | 977.0 (35.0) | 0.043 | 686.6 (31.2) | 1176.0 (34.7) | 0.035 |
| Missing | 6.3 ( 1.4) | 6.0 ( 1.0) | -0.004 | 29.1 ( 1.8) | 37.0 ( 1.3) | -0.004 | 37.1 ( 1.7) | 43.0 ( 1.3) | -0.004 |
| Asparate aminostransferase, IU/L (%) |  |  |  |  |  |  |  |  |  |
| < 20 | 61.7 (13.6) | 65.0 (10.9) | -0.027 | 139.2 ( 8.5) | 218.0 ( 7.8) | -0.007 | 225.0 (10.2) | 283.0 ( 8.4) | -0.019 |
| 20 - 39 | 218.0 (48.2) | 279.0 (46.9) | -0.013 | 741.6 (45.3) | 1218.0 (43.6) | -0.016 | 1025.4 (46.6) | 1497.0 (44.2) | -0.024 |
| 40 + | 172.9 (38.2) | 251.0 (42.2) | 0.040 | 758.1 (46.3) | 1357.0 (48.6) | 0.023 | 950.4 (43.2) | 1608.0 (47.5) | 0.043 |
| Creatinine, mg/dL (%) |  |  |  |  |  |  |  |  |  |
| < 1.2 | 206.5 (45.6) | 277.0 (46.6) | 0.009 | 700.3 (42.7) | 1324.0 (47.4) | 0.047 | 965.0 (43.9) | 1601.0 (47.3) | 0.034 |
| 1.2 – 1.99 | 179.4 (39.6) | 244.0 (41.0) | 0.014 | 632.4 (38.6) | 1048.0 (37.5) | -0.011 | 861.5 (39.1) | 1292.0 (38.1) | -0.010 |
| 2 + | 66.8 (14.8) | 74.0 (12.4) | -0.023 | 306.2 (18.7) | 421.0 (15.1) | -0.036 | 374.1 (17.0) | 495.0 (14.6) | -0.024 |
| Missing | 0.0 ( 0.0) | 0.0 ( 0.0) | 0.000 | 0.0 ( 0.0) | 0.0 ( 0.0) |  | 0.0 ( 0.0) | 0.0 ( 0.0) |  |
| Fibrosis-4 Index (%) |  |  |  |  |  |  |  |  |  |
| < 1.45 | 96.4 (21.3) | 129.0 (21.7) | 0.004 | 197.6 (12.1) | 411.0 (14.7) | 0.027 | 335.2 (15.2) | 540.0 (15.9) | 0.007 |
| 1.45 – 3.25 | 178.7 (39.5) | 239.0 (40.2) | 0.007 | 692.4 (42.2) | 1260.0 (45.1) | 0.029 | 878.7 (39.9) | 1499.0 (44.2) | 0.043 |
| 3.25 + | 171.1 (37.8) | 220.0 (37.0) | -0.008 | 718.1 (43.8) | 1081.0 (38.7) | -0.051 | 948.1 (43.1) | 1301.0 (38.4) | -0.047 |
| Missing | 6.6 ( 1.5) | 7.0 ( 1.2) | -0.003 | 30.8 ( 1.9) | 41.0 ( 1.5) | -0.004 | 38.7 ( 1.8) | 48.0 ( 1.4) | -0.003 |
| Lactate, mmol/L (%) |  |  |  |  |  |  |  |  |  |
| 1_1.2 | 86.8 (19.2) | 118.0 (19.8) | 0.007 | 292.2 (17.8) | 496.0 (17.8) | -0.001 | 376.5 (17.1) | 614.0 (18.1) | 0.010 |
| 2_1.2LT2 | 121.9 (26.9) | 172.0 (28.9) | 0.020 | 476.3 (29.1) | 930.0 (33.3) | 0.042 | 667.0 (30.3) | 1102.0 (32.5) | 0.022 |
| 3_GE2 | 64.8 (14.3) | 96.0 (16.1) | 0.018 | 239.8 (14.6) | 413.0 (14.8) | 0.002 | 327.2 (14.9) | 509.0 (15.0) | 0.002 |
| Missing | 179.2 (39.6) | 209.0 (35.1) | -0.044 | 630.7 (38.5) | 954.0 (34.2) | -0.043 | 830.0 (37.7) | 1163.0 (34.3) | -0.034 |
| Platelet count per microL (%) |  |  |  |  |  |  |  |  |  |
| 150 or higher | 302.6 (66.8) | 401.0 (67.4) | 0.006 | 1046.9 (63.9) | 1893.0 (67.8) | 0.039 | 1391.4 (63.2) | 2294.0 (67.7) | 0.045 |
| < 150 | 150.1 (33.2) | 194.0 (32.6) | -0.006 | 590.1 (36.0) | 897.0 (32.1) | -0.039 | 807.6 (36.7) | 1091.0 (32.2) | -0.045 |
| Missing | 0.0 ( 0.0) | 0.0 ( 0.0) | 0.000 | 1.9 ( 0.1) | 3.0 ( 0.1) | 0.000 | 1.6 ( 0.1) | 3.0 ( 0.1) | 0.000 |
| Total bilirubin, mg/dL (%) |  |  |  |  |  |  |  |  |  |
| < 1 | 347.4 (76.7) | 456.0 (76.6) | -0.001 | 1272.7 (77.7) | 2089.0 (74.8) | -0.029 | 1653.3 (75.1) | 2545.0 (75.1) | 0.000 |
| 1 - 1.2 | 36.7 ( 8.1) | 49.0 ( 8.2) | 0.001 | 136.0 ( 8.3) | 278.0 (10.0) | 0.017 | 203.9 ( 9.3) | 327.0 ( 9.7) | 0.004 |
| 1.2 + | 64.2 (14.2) | 86.0 (14.5) | 0.003 | 202.5 (12.4) | 393.0 (14.1) | 0.017 | 311.5 (14.2) | 479.0 (14.1) | 0.000 |
| Missing | 4.4 ( 1.0) | 4.0 ( 0.7) | -0.003 | 27.7 ( 1.7) | 33.0 ( 1.2) | -0.005 | 32.1 ( 1.5) | 37.0 ( 1.1) | -0.004 |
| White Blood Count per microL (%) |  |  |  |  |  |  |  |  |  |
| 4-10 | 228.8 (50.5) | 274.0 (46.1) | -0.045 | 835.1 (51.0) | 1261.0 (45.1) | -0.058 | 1089.7 (49.5) | 1535.0 (45.3) | -0.042 |
| <4 | 136.0 (30.0) | 202.0 (33.9) | 0.039 | 508.8 (31.0) | 821.0 (29.4) | -0.016 | 692.9 (31.5) | 1023.0 (30.2) | -0.013 |
| >10 | 87.9 (19.4) | 119.0 (20.0) | 0.006 | 295.1 (18.0) | 711.0 (25.5) | 0.075 | 418.1 (19.0) | 830.0 (24.5) | 0.055 |
| C-reactive protein measured (%) | 313.0 (69.1) | 415.0 (69.7) | 0.006 | 1016.3 (62.0) | 1828.0 (65.4) | 0.034 | 1404.8 (63.8) | 2243.0 (66.2) | 0.024 |
| D-dimer measured (%) | 365.9 (80.8) | 486.0 (81.7) | 0.009 | 1366.2 (83.4) | 2354.0 (84.3) | 0.009 | 1817.9 (82.6) | 2840.0 (83.8) | 0.012 |
| **Vital Signs** |  |  |  |  |  |  |  |  |  |
| Highest Temperature (F) (%) |  |  |  |  |  |  |  |  |  |
| < 99 | 156.4 (34.6) | 198.0 (33.3) | -0.013 | 416.6 (25.4) | 816.0 (29.2) | 0.038 | 633.9 (28.8) | 1014.0 (29.9) | 0.011 |
| 99 - 100 | 114.2 (25.2) | 140.0 (23.5) | -0.017 | 325.2 (19.8) | 644.0 (23.1) | 0.032 | 491.0 (22.3) | 784.0 (23.1) | 0.008 |
| 100 - 102 | 126.3 (27.9) | 183.0 (30.8) | 0.029 | 562.0 (34.3) | 878.0 (31.4) | -0.029 | 695.5 (31.6) | 1061.0 (31.3) | -0.003 |
| 102 + | 52.9 (11.7) | 71.0 (11.9) | 0.002 | 327.2 (20.0) | 438.0 (15.7) | -0.043 | 368.4 (16.7) | 509.0 (15.0) | -0.017 |
| Missing | 2.8 ( 0.6) | 3.0 ( 0.5) | -0.001 | 7.9 ( 0.5) | 17.0 ( 0.6) | 0.001 | 11.9 ( 0.5) | 20.0 ( 0.6) | 0.001 |
| Mean Arterial Pressure, mmHg (%) |  |  |  |  |  |  |  |  |  |
| < 60 | 9.8 ( 2.2) | 9.0 ( 1.5) | -0.006 | 24.9 ( 1.5) | 46.0 ( 1.6) | 0.001 | 44.0 ( 2.0) | 55.0 ( 1.6) | -0.004 |
| 60 – 69 | 50.0 (11.1) | 51.0 ( 8.6) | -0.025 | 245.4 (15.0) | 341.0 (12.2) | -0.028 | 289.4 (13.2) | 392.0 (11.6) | -0.016 |
| 70 – 89 | 300.5 (66.4) | 401.0 (67.4) | 0.010 | 1106.5 (67.5) | 1906.0 (68.2) | 0.007 | 1477.4 (67.1) | 2307.0 (68.1) | 0.010 |
| 90 + | 91.8 (20.3) | 133.0 (22.4) | 0.021 | 254.6 (15.5) | 486.0 (17.4) | 0.019 | 386.6 (17.6) | 619.0 (18.3) | 0.007 |
| Missing | 0.6 ( 0.1) | 1.0 ( 0.2) | 0.000 | 7.5 ( 0.5) | 14.0 ( 0.5) | 0.000 | 3.3 ( 0.1) | 15.0 ( 0.4) | 0.003 |
| Lowest Oxygen Saturation (%) |  |  |  |  |  |  |  |  |  |
| < 88 | 6.7 ( 1.5) | 11.0 ( 1.8) | 0.004 | 175.8 (10.7) | 338.0 (12.1) | 0.014 | 179.7 ( 8.2) | 349.0 (10.3) | 0.021 |
| 88 - 92 | 170.9 (37.7) | 263.0 (44.2) | 0.065 | 950.6 (58.0) | 1686.0 (60.4) | 0.024 | 1157.8 (52.6) | 1949.0 (57.5) | 0.049 |
| 93 - 95 | 202.5 (44.7) | 238.0 (40.0) | -0.047 | 419.4 (25.6) | 580.0 (20.8) | -0.048 | 674.2 (30.6) | 818.0 (24.1) | -0.065 |
| 96 + | 59.1 (13.1) | 70.0 (11.8) | -0.013 | 65.2 ( 4.0) | 119.0 ( 4.3) | 0.003 | 148.4 ( 6.7) | 189.0 ( 5.6) | -0.012 |
| Missing | 13.6 ( 3.0) | 13.0 ( 2.2) | -0.008 | 28.0 ( 1.7) | 70.0 ( 2.5) | 0.008 | 40.6 ( 1.8) | 83.0 ( 2.4) | 0.006 |

**Supplemental Table 2. Sensitivity analyses estimating the average treatment effect in the treated population (ATT) in weighted Cox proportional hazards models for the association of corticosteroids with all-cause 90-day mortality among those not on IRS**

|  | **No oxygen supplementation** | **Nasal cannula** | **Combined group: no oxygen plus NC** |
| --- | --- | --- | --- |
|  | **HR (95% CI)** | **HR (95% CI)** | **HR (95% CI)** |
| **Primary analysis** | 1.77 (1.25-2.50) | 1.26 (0.92-1.74) | 1.60 (1.24-2.08) |
| **Subgroup analyses** |  |  |  |
| Restricted to dexamethasone | 1.55 (0.95-2.52) | 1.28 (0.84-1.94) | 1.49 (1.02-2.18) |
| Excluding patients admitted to ICU in initial 48 hours | 1.62 (1.05-2.49) | 1.33 (0.88-1.99) | 1.60 (1.15-2.24) |
| Restricted to patients age 70 and older | 1.69 (1.13-2.54) | 1.49 (0.99-2.24) | 1.72 (1.24-2.40) |
| Models present the ATT (average treatment effect in treated population).  CI = confidence interval  HR = hazard ratio  IRS = intensive respiratory support | | | |

**Supplemental Table 3. Sensitivity analyses with unweighted, multivariable Cox proportional hazards regression models for the association of corticosteroids with all-cause 90-day mortality**

|  | **No oxygen supplementation** | **Nasal cannula** | **Combined group: no oxygen plus NC** |
| --- | --- | --- | --- |
|  | **HR (95% CI)** | **HR (95% CI)** | **HR (95% CI)** |
| **Primary analysis** | 1.87 (1.36-2.57) | 1.39 (1.10-1.76) | 1.66 (1.39-1.99) |
| **Subgroup analyses** |  |  |  |
| Restricted to dexamethasone | 1.87 (1.32-2.66) | 1.37 (1.08-1.74) | 1.66 (1.38-1.99) |
| Excluding patients admitted to ICU in initial 48 hours | 1.85 (1.31-2.62) | 1.31 (1.01-1.69) | 1.63 (1.34-1.98) |
| Restricted to patients age 70 and older | 2.02 (1.40-2.89) | 1.41 (1.09-1.82) | 1.66 (1.36-2.02) |
| CI = confidence interval  HR = hazard ratio  IRS = intensive respiratory support | | | |

**Supplemental Table 4. STROBE Checklist of items for cohort studies**

|  | Item No | Recommendation | Section/Paragraph |
| --- | --- | --- | --- |
| **Title and abstract** | 1 | (*a*) Indicate the study’s design with a commonly used term in the title or the abstract | Abstract: Methods |
|  |  | (*b*) Provide in the abstract an informative and balanced summary of what was done and what was found | Abstract: Methods, Findings, Interpretation |
| Introduction | | |  |
| Background/rationale | 2 | Explain the scientific background and rationale for the investigation being reported | Introduction: Paragraphs 1-2 |
| Objectives | 3 | State specific objectives, including any prespecified hypotheses | Introduction: Paragraph 3 |
| Methods | | |  |
| Study design | 4 | Present key elements of study design early in the paper | Methods: Paragraph 1 |
| Setting | 5 | Describe the setting, locations, and relevant dates, including periods of recruitment, exposure, follow-up, and data collection | Methods: Paragraph 1 |
| Participants | 6 | (*a*) Give the eligibility criteria, and the sources and methods of selection of participants. Describe methods of follow-up | Methods: Paragraphs 1-2 |
|  |  | (*b*) For matched studies, give matching criteria and number of exposed and unexposed | N/A |
| Variables | 7 | Clearly define all outcomes, exposures, predictors, potential confounders, and effect modifiers. Give diagnostic criteria, if applicable | Methods: Paragraphs 3-6 |
| Data sources/ measurement | 8* | For each variable of interest, give sources of data and details of methods of assessment (measurement). Describe comparability of assessment methods if there is more than one group | Methods: Paragraphs 3-6 |
| Bias | 9 | Describe any efforts to address potential sources of bias | Methods: Paragraphs 7-8 |
| Study size | 10 | Explain how the study size was arrived at | All hospitalized COVID+ patients with at least 48h stay, concatenating length of stay to include emergency department/ observational status |
| Quantitative variables | 11 | Explain how quantitative variables were handled in the analyses. If applicable, describe which groupings were chosen and why | Methods: paragraphs 4, 6, Table 2, Supplemental Table 1 |
| Statistical methods | 12 | (*a*) Describe all statistical methods, including those used to control for confounding | Methods: paragraphs 7, 8 |
|  |  | (*b*) Describe any methods used to examine subgroups and interactions | Methods: paragraph 9 |
|  |  | (*c*) Explain how missing data were addressed | Methods: paragraph 8, Table 2, Supplemental Table 1 |
|  |  | (*d*) If applicable, explain how loss to follow-up was addressed | N/A |
|  |  | (*e*) Describe any sensitivity analyses | Methods: paragraph 9 |
| Results | | |  |
| Participants | 13* | (a) Report numbers of individuals at each stage of study—eg numbers potentially eligible, examined for eligibility, confirmed eligible, included in the study, completing follow-up, and analysed | Methods: paragraph 1-2  Results: paragraph 1  Figure 1 |
|  |  | (b) Give reasons for non-participation at each stage | Methods: paragraph 2  Figure 1 |
|  |  | (c) Consider use of a flow diagram | Figure 1 |
| Descriptive data | 14* | (a) Give characteristics of study participants (eg demographic, clinical, social) and information on exposures and potential confounders | Results: paragraphs 1-2  Table 1 |
|  |  | (b) Indicate number of participants with missing data for each variable of interest | Table 2, Supplemental Table 1 |
|  |  | (c) Summarise follow-up time (eg, average and total amount) | Figure 2 |
| Outcome data | 15* | Report numbers of outcome events or summary measures over time | Results: paragraph 2  Table 1, Figure 2, Figure 3 |
| Main results | 16 | (*a*) Give unadjusted estimates and, if applicable, confounder-adjusted estimates and their precision (eg, 95% confidence interval). Make clear which confounders were adjusted for and why they were included | Results: paragraph 2  Results: paragraphs 4, 5  Table 3 |
|  |  | (*b*) Report category boundaries when continuous variables were categorized | Table 2 and Supplemental Table 1 |
|  |  | (*c*) If relevant, consider translating estimates of relative risk into absolute risk for a meaningful time period |  |
| Other analyses | 17 | Report other analyses done—eg analyses of subgroups and interactions, and sensitivity analyses | Results: paragraph 6 |
| Discussion | | |  |
| Key results | 18 | Summarise key results with reference to study objectives | Discussion: paragraph 1-2 |
| Limitations | 19 | Discuss limitations of the study, taking into account sources of potential bias or imprecision. Discuss both direction and magnitude of any potential bias | Discussion: paragraph 7 |
| Interpretation | 20 | Give a cautious overall interpretation of results considering objectives, limitations, multiplicity of analyses, results from similar studies, and other relevant evidence | Discussion: paragraphs 1, 2, 3, 5, 6 |
| Generalisability | 21 | Discuss the generalisability (external validity) of the study results | Discussion: paragraph 3, 4, 5, 6 |
| Other information | | |  |
| Funding | 22 | Give the source of funding and the role of the funders for the present study and, if applicable, for the original study on which the present article is based | Metadata |

*Give information separately for exposed and unexposed groups.
